# Efficacy and Best Practices of Health-care worker Smoking Cessation Treatment in Sub-Saharan Africa

**DOI:** 10.1101/2024.12.08.24318613

**Authors:** W. Davison, M. Sime, W. Khan, E. Yamoah, K. Bhurji, R. Surti

**Affiliations:** University of leeds

## Abstract

**Background:** Tobacco smoking causes over 8 million deaths annually worldwide and is expected to increase by 148% in sub-Saharan Africa by 2030. This puts significant strain on already heavily burdened healthcare systems. Healthcare workers (HCWs) are at the forefront of patient care and play a crucial role in smoking cessation (SC) efforts.

**Objectives:** To assesses the effectiveness and identify barriers to smoking cessation provision by HCWs in sub-Saharan Africa.

**Method:** A systematic review was conducted in accordance with PRISMA guidelines to retrieve studies relevant to the implementation of SC strategies in sub-Saharan Africa and research on the barriers and facilitators of the adoption of SC practices. Studies were retrieved from PubMed, Medline, Ovid, Cochrane Library and Scopus; all included literature was published in English after 2014. Risk of bias and methodological quality were evaluated through the Critical Appraisal Skills Programme (CASP) tool.

**Results:** Twelve studies met the inclusion criteria. HCWs in sub-Saharan with formal training were more likely to offer SC interventions, with lack of training and resources identified as key barriers. Greater HCW engagement with patients also facilitated SC efforts. Inadequate resources, cultural differences and structural failures were further barriers. Socioeconomic and educational differences also influenced quit attempts, with wealthier and better-educated individuals more likely to quit.

**Discussion:** HCW competency and training significantly impacts SC efforts, suggesting the need for comprehensive programs to boost HCW skills and knowledge. Socioeconomic and cultural factors also affected SC outcomes, highlighting the need for tailored health campaigns.

However, the heterogenicity of the evidence base makes it challenging to compare SC interventions and determine if identified barriers are only region specific or generalisable.

**Conclusion:** HCW training and support is crucial for SC provision across sub-Saharan Africa, but structural barriers and sociocultural challenges must be addressed for these programs to succeed. Interventions should therefore both empower HCWs and be tailored to the local area.

## Background

If effective tobacco control strategies are not implemented, the number of smokers in sub-Saharan Africa is estimated to increase to 208 million (148%) by 2030 (Mbongwe and Tapera, 2021). Tobacco smoking is already the leading cause of preventable death worldwide, with over 8 million people dying each year; this includes 1.3 million from second-hand smoke exposure (World Health, 2023). Numerous non-communicable diseases (NCDs) are associated with tobacco use, including chronic respiratory conditions, cardiovascular diseases, cancers, and foetal malformations (West, 2017). Smoking also exacerbates the progression and consequences of communicable diseases, most notably tuberculosis (TB) and Human Immunodeficiency Virus (HIV), making it a critical public health issue in TB and HIV endemic regions. Globally, 60% of people living with HIV/Acquired Immunodeficiency Syndrome (AIDS) are from sub-Saharan Africa, and it remains devastated by the epidemic (Siddiqi, 2019). In this region, where healthcare systems are already strained by both the burdens of NCDs and infectious diseases, the rising prevalence of smoking amplifies the challenge (Agyei and Kumah, 2024). This makes smoking cessation an essential public health priority and a topic that needs exploration.

Tobacco smoking imposes a substantial economic burden, particularly in low-resource settings, such as those in sub-Saharan Africa. Associated diseases not only increase healthcare expenditure at the national level but also impose a heavy financial burden on individuals, as out-of-pocket (OOP) payments account for more than 37% of health spending across Africa (Karamagi et al., 2023). Smoking also reduces productivity, exacerbating poverty in affected communities (Ekpu and Brown, 2015). The cost of treating smoking-related diseases (such as chronic obstructive pulmonary disease (COPD) and lung cancers caused by tobacco use takes a significant portion of health resources that could be used in other medical services (e.g. vaccination programmes) - in South Africa alone, the economic cost of smoking related disease was around 1% of GDP (Boachie et al., 2021). Reducing smoking prevalence through effective cessation programmes would alleviate some of the economic burden on healthcare systems and reallocate resources to areas of greater need for disease prevention.

Healthcare workers (HCWs) can be at the forefront of tobacco cessation efforts. Their regular communication with patients makes them an ideal source to provide smoking cessation advice, offer behavioural support, and provide motivation towards their preparation to quit (La Torre et al., 2020). Evidence from high-income countries shows that brief health provider interventions, such as advice to quit or motivational interviewing, even of short duration over weeks or months, have been associated with a higher rate of successful smoking cessation ((Fiore, 2008; General et al., 2020). However, the effectiveness of healthcare worker-led smoking cessation interventions remains underexplored in sub-Saharan Africa, particularly in resource-limited settings with barriers such as inadequate training, cultural influences, and lack of smoking cessation resources.

HCWs in such settings are often burdened by a high patient-to-worker ratio and lack access to smoking cessation tools, such as nicotine replacement therapies and behavioural counselling programmes (Shangase et al., 2017). There are also significant cultural and societal influences on smoking behaviour within sub-Saharan Africa that must be considered. In some communities, smoking is associated with masculinity, making it harder to disrupt these societal norms (Kodriati et al., 2018). Smoking is also a stigmatised behaviour among many women, leading to the hidden use of tobacco, further complicating efforts to provide cessation support (Thirlway et al., 2021).

The primary objective of this systematic review is to evaluate the effectiveness of smoking cessation interventions delivered by HCWs within sub-Saharan Africa. Furthermore, the review will provide evidence-based recommendations for smoking cessation intervention, which may be integrated into broader healthcare programmes, such as TB-tobacco reduction programmes. This review aims to synthesise the available evidence and to advise policies and practices, reducing the smoking disease burden and improving health outcomes in sub-Saharan Africa.

## Methods

A systematic review was conducted to synthesise the published literature relating to smoking cessation interventions implemented by healthcare workers in sub-Saharan Africa.

This systematic review was reported according to the Preferred Reporting Items for Systematic reviews and Meta-Analyses (PRISMA) which can be found in (Appendix 1) (Page et al., 2021).

### Software

Rayyan systematic review software was used to conduct this review (Ouzzani et al., 2016).

### Pilot Search

A pilot search was conducted by all reviewers to refine the final search strategy by identifying appropriate databases and key terms to include in the search. The pilot search was part of the search validation procedure to ensure that all relevant papers were identified by the search strategy.

This review did not conduct repetitions of the search because the pilot search identified that the literature in this area is not expanding rapidly enough to justify repeated searches.

### Search strategy

This systematic review was conducted across five databases: PubMed, Medline, Ovid, Cochrane Library and Scopus. These databases were used as during the pilot search the majority of relevant articles identified originated from these databases. Furthermore, the academic standard of these databases is high, adding to the reliability of the review, and using several databases ensured that the search conducted in this review was comprehensive.

The query strings used in the search strategy can be found in Appendix 2.

Each reviewer was responsible for conducting a search on one of the databases. All the identified papers were uploaded in the research tool Rayyan which aided in screening.

### Eligibility Criteria

The Population, Intervention, Comparison, and Outcome (PICO) framework was used to establish the inclusion and exclusion criteria of this review (Huang et al., 2006). The inclusion and exclusion criteria are summarised in Table 1. We limited the search of the literature to publications from the last 20 years to ensures the review captures and focuses on contemporary practices and interventions for smoking cessation.

| Inclusion | Exclusion |
| --- | --- |
| **Population:**<br>Studies involving healthcare providers (e.g., doctors, nurses) in any setting.<br>**Intervention:**<br>Research focused on implementing *effective, safe, and cost-effective* smoking cessation treatments.<br>Interventions that address barriers and facilitators to the adoption of smoking cessation practices by healthcare providers.<br>**Study Type:**<br>Implementation studies, RCTs, systematic reviews, and relevant qualitative research<br>Published literature<br>**Outcomes:**<br>Studies reporting on implementation success, adherence to guidelines, and cost-effectiveness.<br>**Setting:**<br>Studies conducted in healthcare environments<br>Studies conducted in sub-Saharan Africa<br>**Language:**<br>English or available English translations.<br>**Publication Date:**<br>Studies published after 2014 | **Population:**<br>Studies not involving healthcare providers.<br>**Intervention:**<br>Research that does not involve healthcare providers or relevant stakeholders in the implementation process.<br>**Study Type:**<br>Case reports, opinion pieces, or studies.<br>Grey literature<br>**Outcomes:**<br>Studies not addressing implementation or provider-level outcomes.<br>**Setting:**<br>Studies that do not provide insights applicable to healthcare settings, such as those conducted in non-healthcare environments.<br>Studies not relevant to Saharan Africa<br>**Language:**<br>Non-English without translation.<br>Studies where the full text is unavailable.<br>**Publication Date:**<br>Studies published before 2014, unless foundational. |

### Screening

All the papers identified in the search were collated in Rayyan and duplicates removed. Papers were initially screened by title, then by abstract and keywords by two independent reviewers (EY+MS and WK+KB) in the research team. They were blinded to everything other than the paper’s title, abstract and keywords, as well as to each other’s decisions. The software identified any disagreements between the reviewers; when discrepancies occurred, the reviewers met to discuss their reasoning behind their decisions and re-reviewed the inclusion and exclusion criteria to decide whether the paper was eligible for the next stage of the screening process. If a consensus could not be reached, disagreements were escalated to a senior reviewer (WD).

Full text screening was carried out on the remaining literature by three independent researchers who were blinded to the others’ decisions. Again, any disagreements identified involved re-examining the eligibility criteria and a discussion between the reviewers until a consensus was reached.

### Extraction

Relevant data was extracted from the final papers included in the review into a prepared excel document. The following data points were extracted: year of publication, journal of publication, authors, study deign, intervention details, study outcomes, study conclusions, contextual factors and setting, and risk of bias information.

The results are summarised in a table in the results section.

### Quality Assessment

The Critical Appraisal Skills Programme (CASP) tool was used to assess the methodological quality and the risk of bias of the studies included in this review. The CASP tool was adapted to address the quality of methods for verifying smoking abstinence, intervention type, and socioeconomic and age variation within the sample. Each paper was assessed by two independent reviewers and the overall quality was categorised as either low, medium or high. Disagreements between reviewers were addressed through discussion and third reviewer if necessary.

## Results

This review found 12 studies that met the inclusion criteria (Figure 1). Of these studies, 3 were qualitative, involving interviews, and 1 paper was a mixed-method study involving both interviews and a questionnaire. Of the remaining 8 papers, all these were quantitative, including 2 randomised control trials (RTCs), 1 non-randomised control trials (RTCs) and 5 cross sectional studies. 2 papers were determined as low evidence, 7 as medium, and 3 as high

**Figure 1.**
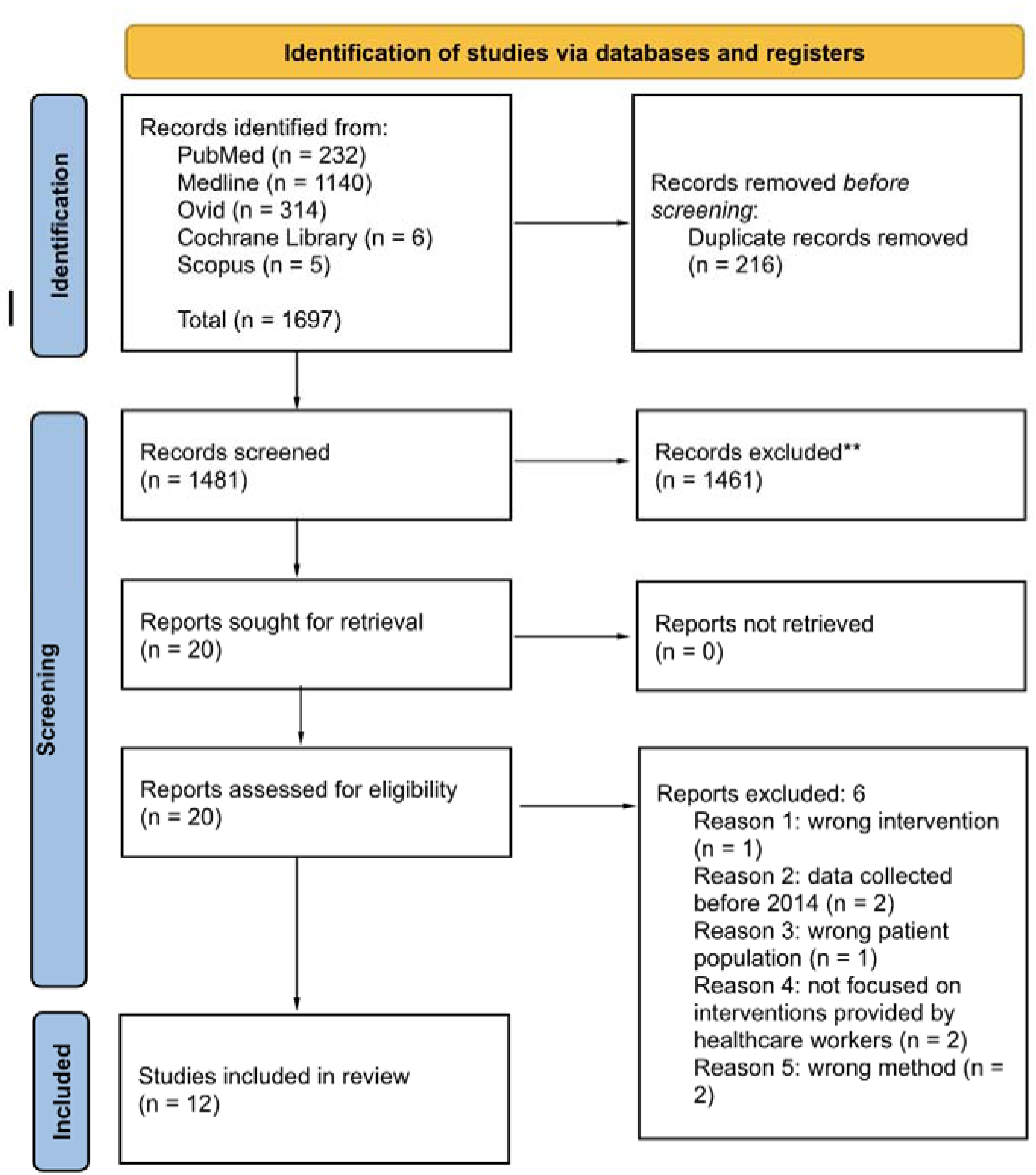
RISMA flow diagram showing the number of papers included in the identification, screening, and extraction process of this review

**Table 2.** study characteristics.

| Study (country) | Study Type | Intervention | Summary of findings | Quality appraisal |
| --- | --- | --- | --- | --- |
| (Bokoro et al., 2020) South Africa | cross-sectional quantitative | Study targeted tobacco users at an outpatient department in Krugersdorp, South Africa (May–August 2016). A total of 275 participants were recruited using consecutive sampling. Nurses identified tobacco users, who were then briefed, consented, and surveyed using a researcher-administered questionnaire on their tobacco use and quit attempts. Data were analyzed using descriptive statistics and logistic regression to explore factors associated with quitting. | Quit attempts: 74% reported ever making a quit attempt, with 55% attempting in the last year. Motivation: Health concerns were the primary motivation for quitting. Barriers: Stress and cravings were the most common barriers. Factors Associated with Quit Attempts - Marital status: Married participants were more likely to attempt quitting (OR: 2.13). Other factors: White population group and employment status were not significantly associated with quit attempts. | Medium |
| (Du Plooy et al., 2016) South Africa | Cross sectional observational quantitative | Participants were recruited consecutively over a 2-month period in 2016, with eligibility based on age (18-59 years), clinical stability, and literacy. Data were collected through medical records and a structured interview using a modified version of the Global Adult Tobacco Survey (GATS), the Fagerström Test for Nicotine Dependence (FTND), and the Decisional Balance for Cigarette Smoking (DBCS). | Most daily smokers consumed $\leq 20$ cigarettes per day, and 55.7% had high or very high nicotine dependence. (71.7%) perceived more positives than negatives about smoking, while 23.6% saw more negatives, and 4.7% were neutral. 59.4% of smokers had attempted to quit within the last year, with non-daily smokers more likely to have tried quitting (84.2%) compared to daily smokers (54%). Only 43.4% had received advice to quit smoking from a health professional in the past year. | High |
| (Louwagie, G.M. et al., 2014) South Africa | Randomised control trial | Conducted in six TB clinics in Soshanguve, South Africa, aimed to assess the impact of brief motivational interviewing (MI) on smoking cessation among TB patients. Adults with TB were | Self-Reported 6-Month Abstinence: Higher in MI group (21.5%) vs. control (9.3%), RR = 2.29 (95% CI 1.34–3.92). Biochemically Verified Abstinence: MI group (28.9%) vs. control (13.3%), crude RR = 2.21 | Medium |
|  |  | recruited who were current smokers (excluding those under 18, too ill, non-smokers, or on TB treatment for over a month). Participants were randomly assigned to either an intervention group (n=205) (brief MI session) or a control group (n=204). Both received standardised CS message from tb Nurse. Intervention group also received a brief MI. Self-reported smoking abstinence was measured. Primary outcome was 6-month sustained abstinence; secondary outcomes included 3-month abstinence, 7-day point-prevalence abstinence, and quit attempts.<br>Relative risks and absolute risk differences were calculated using Poisson regression. Primary analysis followed intention-to-treat principles. | (95% CI 1.08–4.51); adjusted RR = 2.33 (95% CI 1.11–4.90). Overall Verification (all participants): MI group (11.7%) vs. control (5.4%), RR = 2.15 (95% CI 1.06–4.40). Self-Reported 3-Month Abstinence higher in MI group (25.4%) vs. control (12.8%), RR = 1.98 (95% CI 1.24–3.18); biochemically verified higher in MI group. Self-Reported 1-Month Abstinence 7-day point prevalence higher in MI group (35.1%) vs. control (22.1%), RR = 1.59 (95% CI 1.10–2.31); biochemically verified rates also higher in MI group. Intervention Details: 68.1% Advised to stop smoking, 10.3% received TB-related cessation message |  |
| (Louwagie et al., 2022) South Africa | randomised controlled trial (RCT) | Participants: Adults (≥18 years) with drug-sensitive pulmonary tuberculosis (PTB), either starting or within one month of TB treatment, and who were tobacco smokers or harmful drinkers. 574 participants took part (291 control, 283 intervention) out of 2099 screened. Pro life intervention: three motivational interviewing (MI) sessions (15-20 minutes each), spaced one month apart, delivered by trained lay | No significant difference between the two groups - The rate of successful TB treatment was 70.1% in the control arm and 67.8% in the intervention arm, with an odds ratio (OR) of 0.90 (95% CI 0.64 to 1.27), Similarly, cure rates were 39.9% in the control arm and 43.6% in the intervention arm, with an OR of 1.16 (95% CI 0.83 to 1.63), again showing no significant impact of the intervention.<br>The secondary outcomes also did not show significant differences. | Medium |
|  |  | health workers (LHWs), as well as bi-weekly SMS messages over 12 weeks, focusing on TB treatment adherence, smoking cessation, and alcohol reduction.<br>Control: Usual care as per local TB treatment protocols<br>Primary outcome was TB treatment success at 6-9 months, secondary outcomes included sputum conversion, smoking abstinence, changes in alcohol consumption, and ART adherence. | Continuous smoking abstinence rates at 6 months were 6.4% in the intervention arm versus 7.2% in the control arm (OR 0.76, 95% CI 0.35 to 1.63). Alcohol use reduction, as measured by the AUDIT score, showed only minimal changes, with a mean reduction of 0.04 points in the intervention arm compared to a 0.55-point increase in the control arm at 6 months. |  |
| (Mahoto et al., 2023) Zambezi region, Namibia | Mixed-method | The aim was to identify the barriers that prevent healthcare workers from providing supportive care services to patients in the Zambezi region of Namibia. The information was gathered from 129 respondents who were HCWs who had lived in one of the 8 constituencies of the Zambezi region in Namibia for over 5 years and aged between 17 and 60.<br>The qualitative stage: involved focus group discussions to identify perceived barriers to smoking cessation. These were completed until saturation was maintained. Then used thematic analysis. The quantitative stage: involved cross sectional survey on knowledge, attitude, and practices regarding smoking cessation | Knowledge and Training Deficits were identified - Inadequate Training: 61.7% of HCWs cited lack of training as a major barrier to providing effective smoking cessation (SC) services. Insufficient Knowledge: 41.5% of HCWs felt their knowledge of SC interventions was insufficient. Certain Attitudes were also identified as hindering smoking cessation. These included - Perception of Responsibility: HCWs often viewed SC as outside their primary duties, typically stopping at basic advice. Cultural Barriers: HCWs hesitated to discuss SC due to cultural norms, especially with older or male patients. Role Misconceptions: Some nurses felt undervalued in their role in SC, leading to frustration.<br>There were also practical barriers reported such as time Constraints: 66% of HCWs reported that lack of time during consultations hindered SC efforts, lack of Resources: 60.6% of HCWs cited the absence of SC guidelines, and 56.4% noted a lack of educational<br>resources for patients. Also Patient-Related Challenges<br>- Low Patient Interest: 45.7% of patients were disinterested in SC, and 63.8% struggled with adherence. Structural Issues were reported: Difficulties in patient follow-up due to logistical barriers like incorrect contact information or relocation. | Medium |
| (Odukoya, O.O. et al., 2014)<br>Nigeria | Non-Randomised control trial | 6 co-educational secondary schools, 3 control, 3 intervention, were selected using multi-stage sampling. Exclusions were made for schools with prior anti-smoking programs and first-year students. N=503 total students for control and n=503 for intervention schools. Data collection included pre- and post-intervention questionnaires assessing smoking-related knowledge, attitudes, and practices. The intervention, lasting three months, featured health talks, leaflets, and posters. | Pre-intervention questionnaires: 973 collected, response rates of 93.6% for intervention group and 95.2% for control group. Post intervention: 948 questionnaires were collected, with response rates of 91.3% for the intervention group and 92.5% for the control group. The mean age of respondents was 14.2 years, with 94.1% under 18 and 52.5% were female.<br>Before intervention, knowledge scores were similar between the groups -intervention: 8.39 control: 8.41 (scores are calculated based off a questionnaire containing 16 knowledge, 7 attitude and 7 practice items) After intervention, the intervention group's knowledge score significantly increased to 11.98 ($p<0.001$), whereas the control group did not show significant change. Attitudes in the intervention group improved significantly, from a mean score of 13.24 to 14.99 ($p<0.001$), compared to insignificant in control. Smoking behaviour remained stable in the intervention group, with a 1% decrease in those smoking in the last 30 days, compared to a 0.5% increase in the control group. | Medium |
| | | | The desire to quit among current smokers in the intervention group rose significantly from 47.4% to 85.7% ($p=0.029$), compared to control 46.7% pre-intervention and 47.1% post-intervention (not significant: $p= 0.979$) | |
| (Odukoya, O. et al., 2017) Nigeria | Cross-sectional observational quantitative | cross-sectional descriptive analysis of the documentation of tobacco use and cessation services provided by physicians in six teaching hospitals across Nigeria. Reviewed 1,588 patient health records from both in-patient and out-patient departments, with systematic sampling used to select ten case files per ward or clinic. Data collection covered a 6-month retrospective period, focusing on patients aged 12 years and older. A checklist was used, focusing on the documentation of tobacco use and cessation services using the 5A's approach. | Only 12.9% of tobacco users received cessation advice, 2.6% had their readiness to quit assessed, and 1.5% were offered assistance to quit. Males were 1.86 times more likely to have their tobacco use documented, in-patients were 2.14 times more likely, and patients with tobacco-related conditions were twice as likely to have their tobacco status recorded. | Low |
| (Rossouw and Filby, 2022) Botswana, Cameroon, Ethiopia, Kenya, Nigeria, Senegal, Tanzania, and Uganda. | cross-sectional quantitative study using secondary data | Analysed data from the Global Adult Tobacco Surveys (GATS). A total of 57,857 individuals participated across the countries included in this study. These were chosen based on GATS data availability. The analysis focused on two tobacco cessation outcomes: (1) former tobacco users and (2) current tobacco users who attempted to quit. Concentration indices were used to measure | Wealthier individuals in Cameroon, Ethiopia, Kenya, and Uganda are more likely to attempt quitting range from 0.100 in Kenya to 0.382 in Ethiopia on the CIE, all statistically significant.) Higher education levels are associated with more quit attempts, with significant disparities in countries like Ethiopia (0.222), Kenya (0.331), and Nigeria (0.151). In Former Tobacco Users (TCF), Education also drives disparities | Medium |
|  |  | wealth and education related inequalities in tobacco cessation, and the Erreygers' Corrected Concentration Index (CIE) and decomposition analysis were applied. | Cameroon (CIE = 0.176, $P < 0.001$), Ethiopia (CIE = 0.199, $P < 0.001$), Kenya (CIE = 0.223, $P < 0.001$), Tanzania (CIE = 0.104, $P = 0.0051$), and Uganda (CIE = 0.109, $P < 0.001$), with urban-rural residence impacting inequalities differently across countries Botswana (31%), Cameroon (56%), Nigeria (22%), and Tanzania (17%) Wealth, education, and urban residence are key factors in tobacco cessation inequalities. Gender also plays a role, with Botswana (18%), Cameroon (27%), Kenya (17%), Tanzania (12%), and Uganda (27%) showing increasing educational inequality with respect to female cessation attempts. Misinformation was significant in Ethiopia (52%) and Nigeria (11%), for explaining inequalities in former smokers, about tobacco health risks contribute to these disparities, |  |
| (Rutebemberwa et al., 2021)<br>Uganda | qualitative exploratory | Qualitative exploratory study using focus group discussions (FGDs) and key informant interviews (KIIs). These occurred in nine health facilities in Uganda, including Kampala, eastern, northern, and western regions. Purposive selection was used FGDs: 81 health workers (27 males, 54 females). KIIs: 8 key informants (4 males, 4 females). Analysed using QDA Miner software. Thematic content analysis with saturation achieved. | Recommendations of improving coordination with organizations like the media, empowering patients through education, ensuring continuous follow-up, addressing gaps in health worker training, and upgrading clinical information systems for better documentation and proactive care. Strengthening the health system, particularly through resource allocation and staff training, was seen as critical for successful integration. | High |
| (Shangase et al., 2017)<br>South Africa | qualitative exploratory | The study was conducted at a TB hospital in KwaZulu- | Participants (average smoking history of 10 years) struggled with | High |
|  |  | Natal, South Africa, involving 20 inpatients with drug-resistant TB (DR-TB) who were smokers, selected through purposive sampling. Data were collected through in-depth interviews in September and October 2015. The interviews, conducted in isiZulu and English, focused on smoking behaviour, knowledge of health risks, interest in quitting, and barriers to cessation. Thematic analysis was used to identify key themes and insights. | nicotine addiction, cravings, and failed quit attempts. Smoking was deeply embedded in daily routines (e.g., after meals), making cessation challenging. Most participants reduced their cigarette consumption but couldn't fully quit. Lack of knowledge about effective quit strategies. Lack of willpower was frequently cited as a significant barrier. Psychosocial stress, including hospital boredom and anxiety, made quitting harder. Peer influence, both in and outside the hospital, contributed to continued smoking. Structural barriers. Health education sessions in the hospital were viewed as ineffective and lacked follow-up interventions. Easy access to cigarettes within the hospital and from visitors discouraged quitting. Motivation for cessation. 18 participants expressed motivation to quit due to their TB diagnosis but were frustrated by the lack of support and resources. Wanted pharmacological and psychological support |  |
| (Tamirat, 2021)<br>Hadiya Zone,<br>Southern<br>Ethiopia | Cross-sectional<br>quantitative | Purposive selection of hospitals with proportional allocation of sample size to each hospital, then simple random sampling within hospitals. A total of 323 HCWs were included. HCWs SC practices assessed using a self-administered questionnaire based on the "5 A's" model (Ask, Advise, Assess, Assist, and Arrange follow-up). | Most HCW did not provide SC interventions to their patients. The practice of all components of the 5A's model was minimal, with most healthcare workers either "sometimes" or "never" engaging in these interventions. The following were significantly associated with a greater likelihood of providing smoking cessation interventions: Male (AOR = 2.25, 95% CI 1.31–6.23), those with better knowledge (AOR = 3.25, 95% CI 1.965–9.332). | Medium |
|  |  |  | those who had received formal training (AOR = 6.5, 95% CI 2.366–11.557, and those with fewer years of service (AOR = 4.75, 95% CI 1.756–9.547). |  |
| (Thirlway et al., 2021) Uganda | Cross sectional qualitative exploratory study | 25 ART patients (19 men, 6 women) aged 28–56 were interviewed. 12 were current tobacco users (10 men, 2 women) and 13 former users (9 men, 4 women) who quit after HIV diagnosis. Also interviewed 13 health workers interviewed from 13 clinics, representing various regions. Convenience sampling was used. Thematic and narrative analyses were performed | Barriers to tobacco cessation were identified - Concealment of Tobacco Use: Many participants hid their tobacco use from health workers, impacting cessation support and data accuracy. Family Support: Positive family support played a crucial role in successful smoking cessation. Social and Economic Factors: Social isolation and economic hardship, including food insecurity, made quitting tobacco more difficult. Gender Roles and Masculinity: Social norms and masculinity influenced smoking behaviours, with sociable masculinity making cessation harder for some men. Additionally, Tobacco use was a barrier to ART adherence, with smoking and drinking complicating adherence efforts. | Low |

### Barriers to smoking cessation

#### Training

Insufficient training was highlighted as a significant barrier by several studies to providing CS therapy. A mixed-method study in Namibia (Mahoto et al., 2023) identified that many (61.7%) HCWs felt that inadequate training hindered their ability to provide SC services. From the patient perspective, patients expressed frustration with the lack of knowledge and effective strategies provided by HCWs in TB hospitals in South Africa (Shangase et al., 2017).

By contrast, effective training facilitated better SC outcomes. In Ethiopia, Tamirat (2021) it was reported that HCWs with formal training were much more likely to offer SC interventions (AOR = 6.5, 95% CI 2.366–11.557) in the form of the “5 A’s” model (Ask, Advise, Assess, Assist, Arrange: these are WHO recommendations for steps that primary care providers can take to assist tobacco users in short consultations). Additionally, empowering HCWs through targeted education was emphasised as a method of improving CS advice and engagement (Louwagie, G.M. et al., 2014; Rutebemberwa et al., 2021).

#### Knowledge

Knowledge of healthcare workers was found to play a role in CS provision. It was reported that 41.5% of sampled HCWs in Namibia felt that they lacked sufficient knowledge of SC interventions (Mahoto et al., 2023). Increased knowledge also significantly positively influenced SC practices, with HCWs having better knowledge being 3.25 times more likely to offer SC interventions in Ethiopia (Tamirat, 2021). Lack of knowledge on the part of the patient was also identified as a barrier to CS for TB patients (Shangase et al., 2017) and misinformation about tobacco health risks contributed to inequalities among former smokers in Ethiopia (52%) and Nigeria (11%) (Rossouw and Filby, 2022)

#### Resources

Additionally, resource limitations were identified as a barrier to CS services. In Namibia 60.6% of HCWs reported a lack of smoking cessation guidelines (Mahoto et al., 2023), and an absence of educational materials (56.4%) as well as time constraints (66%). Patients also felt a lack of lack of support and resources for quitting (Shangase et al., 2017). Furthermore, weak clinical information systems made it difficult to track patient progress and follow-up (Rutebemberwa et al., 2021). Misconceptions about the HCW’s role in SC services was also noted, with HCWs feeling that cessation support was outside their primary duties (Mahoto et al., 2023).

#### Cultural and social factors

Cultural and social factors also played a significant role. Economic disparities were particularly highlighted (Thirlway et al., 2021; Rossouw and Filby, 2022). In a multi-country analysis across eight sub-Saharan African nations (Botswana, Cameroon, Ethiopia, Kenya, Nigeria, Senegal, Tanzania, and Uganda) (Rossouw and Filby, 2022), wealthier individuals were more likely to attempt quitting smoking. In Ethiopia, the Corrected Concentration Index (CIE) for wealth was 0.382, indicating significant inequality in quit attempts based on socioeconomic status. Similar trends were observed in Kenya (CIE = 0.100) and Uganda (CIE = 0.382). Differences in education also drove disparities in Cameroon (CIE = 0.176, P < 0.001), Ethiopia (CIE = 0.199, P < 0.001), Kenya (CIE = 0.223, P < 0.001), Tanzania (CIE = 0.104, P = 0.0051), and Uganda (CIE = 0.109, P < 0.001). Additionally. Urban residents were more likely to quit smoking than rural dwellers in some countries. In Botswana, urban residents accounted for 31% of cessation attempts, in Cameroon 56%, and in Nigeria 22%. Gender also proved a barrier in some situations for smoking cessation. In Uganda, gender roles were identified as a barriers to cessation among people living with HIV (Thirlway et al., 2021). Odukoya, O. et al. (2017) found that males were 1.86 times more likely to have their tobacco use documented. By contrast, Mahoto et al. (2023) reported cultural hesitancy in discussing smoking with male patients. Gender also related to education levels, with a higher concentration of women among less educated groups and is associated with educational inequalities in CS (Rossouw and Filby, 2022).

### Facilitators of Smoking Cessation

As well as addressing training and knowledge deficits, other factors were identified as facilitating better SC outcomes. In Uganda, family encouragement was felt to play an important role in helping patients quit smoking (Thirlway et al., 2021). Introducing motivational interviewing (MI) suggested some promise for increasing rates of CS. The randomized controlled trial conducted by Louwagie, G.M. et al. (2014) in South Africa found that brief MI sessions significantly improved smoking abstinence rates among tuberculosis (TB) patients compared to standard care. The study reported a 6-month self-reported abstinence rate of 21.5% in the MI group, significantly higher than the 9.3% observed in the control group (RR = 1.98, 95% CI: 1.24–3.18). However, a more recent trial by Louwagie et al. (2022), targeting TB patients with a 3 MI program as well as bi-weekly SMS messages, found no significant differences in TB treatment success or smoking abstinence rates between the intervention and control groups.

In Nigeria, a non-randomized trial by Odukoya, O.O. et al. (2014) targeting adolescents involved an educational intervention in six secondary schools. The intervention included health talks, leaflets, and posters aimed at improving smoking-related knowledge, attitudes, and practices. While the study demonstrated significant improvements in smoking knowledge and attitudes among students, it did not result in a significant reduction in smoking prevalence.

Several studies also highlighted structural gaps that need to be addressed for more effective SC interventions. Rutebemberwa et al. (2021) recorded recommendations for better coordination among health facilities, improved documentation systems, and increased resource allocation. Shangase et al. (2017) underscored the importance of addressing structural barriers such as inadequate follow-up systems, and easy access to cigarettes within healthcare settings.

## Discussion

This systematic review evaluates the efficacy of SC interventions delivered by HCWs across sub-Saharan Africa, exploring the strategies employed and identifying gaps in knowledge, attitudes, and behaviours that impede effective SC treatment. Included studies were conducted in a range of countries, such as South Africa, Namibia, Nigeria, Botswana, Cameroon, Kenya, Senegal, Tanzania, and Uganda. Collectively, the studies from 2014 to 2024 provide insight into the current facilitators and barriers shaping SC efforts in sub-Saharan Africa. However, the majority were region-specific and may not reflect broader national trends. An exception to this was Rossouw and Filby (2022), who utilized Global Adult Tobacco Surveys (GATS) across eight countries, providing more generalized data.

Diverse methodologies were used, such as qualitative analyses, observational studies, randomised controlled trials, and mixed methods approaches.

### Role of HCWs

HCWs emerged as pivotal in SC service delivery, with their competency being consistently highlighted as a critical factor. Evidence from Ethiopia (Tamirat, 2021) demonstrated that better-trained HCWs, using the 5As model, were more likely to provide SC support, while interventions aimed at enhancing knowledge and skills increased engagement and advice provision, as seen in 2 studies (Louwagie, G.M. et al., 2014; Rutebemberwa et al., 2021). Conversely, inadequate training negatively impacted the delivery of SC, as noted in Namibia (Mahoto et al., 2023). These findings underscore the need for robust, comprehensive training programs that equip HCWs with the necessary skills to offer effective SC interventions and encourage higher levels of HCW engagement.

Increased patient engagement by HCWs was also linked to improved SC outcomes. For example, Louwagie, G.M. et al. (2014) demonstrated the success of motivational interviewing combined with text messaging in reducing smoking rates among TB patients (RR = 2.33, 95% CI 1.11–4.90). While a later study by Louwagie et al. (2022) showed no significant difference, other research—such as Ohkado et al. (2024) study in the Philippines using the ABC (Ask, Brief advice, Cessation support) model—indicated that increased HCW interaction substantially enhances SC efforts in TB patients. Additionally, Blebil et al. (2014) in Malaysia showed that hands-on approaches, including additional counselling sessions, provided significant advantages for SC success. A systematic review by Stead et al. (2016) also provided further evidence in favour of behavioural support interventions for SC by HCWs. These results emphasize the importance of sustained, cost-effective HCW-patient interactions, with even simple communication methods like phone calls yielding significant improvements.

However, HCWs in sub-Saharan Africa also face numerous structural and practical barriers in delivering SC interventions. Studies such as those by Mahoto et al. (2023), Shangase et al. (2017) and Rutebemberwa et al. (2021) highlighted key challenges, including insufficient resources, a lack of clear guidelines and educational material, and inadequate healthcare funding. Time constraints and lack of training further hindered implementation. The impact of such barriers may help explain the findings from Odukoya, O. et al. (2017) whose audit of Nigerian patient records revealed only 12.9% of users across 6 teaching hospitals in Nigeria received SC advice. The identification of these barriers further highlights the importance of resource allocation and comprehensive training to overcome these barriers and empower HCWs to provide SC services.

### Socioeconomic factors

Demographic and social factors, such as, gender and educational levels, also appeared to influence smoking cessation outcomes. Rossouw and Filby (2022) identified low literacy levels as being linked to poorer smoking cessation outcomes, with women more likely to be at a lower literacy level. When targeting this population with public health campaigns, it is important to consider that low literacy can limit the understanding of smoking-related health risks, reducing the campaigns effectiveness. This was found in Malaysia, where SC services that failed to account for low education levels were perceived as not user-friendly (Chean et al., 2019). This emphasises the need to tailor services to the education levels of the local population. Additionally, socio-cultural acceptance of tobacco use further increases the challenges of smoking cessation efforts. This was noted throughout the papers in this review as well as the wider literature. For instance, Mahoto et al. (2023) reported cultural hesitancy in female HCW in discussing smoking cessation with older male patients, and Thirlway et al. (2021) noted masculinity as a barrier to smoking cessation. This is reflected in the wider literature as well. Chean et al. (2019) discussed how the offer and acceptance of cigarettes between friends and relatives in Malaysia is viewed as a gesture of friendship, and Egbe et al. (2016) discusses the importance of smoking in cultural ceremonies, including weddings, in Nigeria. Other important barriers to effective engagement in SC programmes included: general lack of public health campaigns, stigma surrounding smoking and limited access to healthcare services and nicotine replacement therapies (Shangase et al., 2017).

Economic instability and high unemployment rates can further exacerbate these issues, and continued smoking or relapse amongst those who attempt to quit was noted in those with financial stress (Rossouw and Filby, 2022; Shangase et al., 2017). The relationship with socioeconomic status and health outcomes, especially in marginalised populations, is well established in the literature (British Medical Association, 2017). The association between lower socioeconomic status and health risk behaviours, particularly smoking, may be related to higher levels of stress, poorer health literacy and a lower perception of health messaging (World Health Organization, 2010).

The review also draws attention to the integration of smoking cessation measures into existing health programs, specifically tuberculosis treatment initiatives. Shangase et al. (2017) revealed that HCWs expressed concerns about the low prioritisation of smoking cessation, especially in the context of major health issues like TB and HIV. This raises important questions about how healthcare systems in resource-limited settings can effectively balance the competing demands of numerous health priorities. Interestingly, a more recent study by Rutebemberwa et al. (2021), underscored the potential synergistic advantages by embedding smoking cessation services into TB treatment protocols, which could improve health outcomes for affected populations. A more holistic approach to patient care could be achieved from this integration, allowing both tobacco use and TB to be addressed simultaneously Rutebemberwa et al. (2021). Perhaps a multifaceted approach focused on resource allocation, training, and systemic changes, is required to address these challenges.

### Recommendation for further research

Although some studies, for instance, Bokoro et al. (2020) and Du Plooy et al. (2016), highlight the importance of motivations to quit and quit attempts, there is a lack of sufficient data on the long-term success of cessation programmes. There is also a need for further exploration into effective treatments. Particularly, the conflict in findings between the efficacy of brief motivational interviews for SC in those with tb between Louwagie et al. (2022) and Louwagie, G.M. et al. (2014) prompts the need for further research and evidence. There was a lack of RCTs for smoking cessation interventions specifically in sub-Saharan Africa and further high-quality evidence such as these are needed to draw more definitive conclusions.

Additionally, approaches targeting certain limited demographics, such as youth-targeted anti-smoking campaigns and text-messages interventions for tb patients warrant further exploration into how they can be adapted to target wider populations (Odukoya et al., 2014, Louwage et al., 2022).

Furthermore, additional research that addresses institutional barriers, such as those acknowledged by Mahoto et al. (2023), including the burden of excessive workload or paperwork, the limited availability of smoking cessation products and educational materials for patients, and the lack of smoking cessation guidelines, will also be essential for improving the accessibility and sustainability of these programmes.

Lastly, further research is needed to explore countries individually to assess which smoking cessation strategies are most appropriate for specific contexts. Further to this, exploration into the social and cultural barriers to smoking cessation, such as those previously discussed, will be instrumental in the successful implementation of smoking cessation interventions in individual contexts and so should be prioritised.

### Recommendations

Healthcare workers (HCWs) play a pivotal role in the delivery and success of smoking cessation (SC) interventions. Therefore, comprehensive training for HCWs is essential. This is a challenge in resource limited regions, so priority should be given to promoting structured approaches to SC support, such as the WHO’s 5As model, and equipping HCWs with knowledge of available treatment options like nicotine replacement therapy and behavioural support techniques. It is also crucial to emphasize that SC falls within the professional remit of HCWs, with clear guidance on identifying and engaging smokers.

Additionally, structural barriers like insufficient SC guidelines, inadequate educational materials, and limited time for patient interaction need to be addressed. Policies should focus on resource allocation, improving clinical information systems, and ensuring that HCWs are equipped with the tools they need to provide effective and sustained smoking cessation support.

Tailoring SC interventions to address specific population needs is also important. In more economically deprived areas, there are poorer SC outcomes. However, interventions must account for varying education and literacy levels, ensuring that the content is comprehensible and actionable for all groups. Addressing social and cultural barriers, such as gender dynamics, is equally important. HCWs should be supported in overcoming these challenges through culturally sensitive approaches to SC advice.

Furthermore, integrating tobacco cessation services into existing healthcare frameworks, such as tuberculosis (TB) and HIV care, can make smoking cessation routine in patient management. This strategy would allow for the effective targeting of high-risk populations, ensuring that smoking cessation is addressed alongside other significant health concerns, despite resource limitations.

### Limitations

A primary limitation of this review is the heterogeneity of the evidence base, which complicates direct comparisons between studies. The included research employed a wide variety of methodologies, ranging from cross-sectional surveys to randomized controlled trials, qualitative and quantitative. The outcomes also varied significantly, with some focusing on barriers to SC provision for HCWs, some exploring differences in SC between patient groups, and others exploring interventions. The studies were also across a wide range of demographics, with different cultures, relative education, and wealth. This diversity makes it difficult to evaluate the relative effectiveness of different cessation strategies and whether barriers identified were specific to an area or more generalisable. It also made the synthesise of findings more challenging. There was also a lack of high-quality evidence, which further limited the robustness of the conclusions that could be drawn.

Furthermore, the scope of the review was limited to English-language publications from 2014 onward, which may have excluded key studies conducted in other languages or earlier foundational work. This language and temporal restriction could have omitted important regional data or emerging trends in tobacco cessation that are not widely reported in the English-language literature. As such, our findings may not fully reflect the breadth of available evidence on smoking cessation interventions in sub-Saharan Africa.

Finally, while the focus on sub-Saharan Africa provides valuable insights, the region’s vast diversity is another limitation. Healthcare systems, socio-cultural attitudes, and tobacco use behaviours vary widely between countries, influencing the applicability and success of smoking cessation interventions. Drawing generalisations from each region to across sub-Saharan Africa may obscure important country-specific nuances, limiting the relevance of our recommendations in certain contexts. While the findings did in general agree with the wider literature on SC in lower income countries, one must also be careful about generalising these findings beyond sub-Saharan Africa because of specific cultural factors. Additionally, the inclusion of South Africa, a country classified not classified as low-income, could skew the interpretation of results, as its healthcare infrastructure and socio-economic conditions may not reflect those of lower-income nations in the region and globally.

## Conclusion

This systematic review highlights the critical role that HCWs play in SC efforts across sub-Saharan Africa, while also identifying significant gaps in training, resources, and healthcare infrastructure that hinder the effectiveness of these interventions. Although some strategies, such as the 5As model, motivational interviewing, and integrated approaches with tuberculosis care, have demonstrated potential, structural barriers like inadequate training, limited access to SC guidelines, and sociocultural challenges must be addressed for these programs to succeed. Tailoring interventions to the unique cultural and socioeconomic contexts of different populations is essential to improving SC outcomes.

Moreover, socioeconomic factors, including low literacy rates and cultural attitudes toward tobacco use, further complicate SC efforts. These factors often limit the reach and efficacy of public health campaigns, particularly among marginalised populations, such as women and those with lower educational backgrounds. Cultural norms, such as the social acceptance of smoking in certain contexts, exacerbate the challenge. As such, SC interventions must be adaptable to the unique demographic, social, and cultural contexts of specific populations, ensuring that they are accessible and sensitive to local practices.

While the findings align with broader global trends in low-resource settings, this review also underscores the need for further research, particularly through randomised controlled trials, to establish robust evidence-based practices for the region. Expanding the evidence base on long-term SC outcomes and evaluating country-specific strategies is necessary to refine SC interventions. Additionally, exploring SC strategies that address specific subpopulations, such as youth and women, and understanding the institutional barriers faced by HCWs, are critical areas for future investigation.

In conclusion, smoking cessation in sub-Saharan Africa requires a multifaceted approach that combines comprehensive HCW training, resource allocation, and culturally sensitive interventions. Integrating SC services into existing healthcare frameworks, particularly in resource-limited settings, is a promising strategy for improving health outcomes. However, the success of these efforts depends on addressing the socio-economic, cultural, and structural challenges that impede SC interventions.

## Supporting information

Louwagie et al., 2014 CASP checklist

Tamirat, 2021 CASP checklist

Rossouw and Filby, 2022 CASP checklist

Du Plooy et al., 2016 CASP checklist

Mahoto et al., 2023 CASP checklist

Odukoya et al., 2017 CASP checklist

Odukoya, O.O. et al., 2014 CASP checklist

Shangase et al., 2017 CASP checklist

Bokoro et al., 2020 CASP checklist

Rutebemberwa et al., 2021 CASP checklist

Thirlway et al., 2021 CASP checklist

Louwagie, G.M. et al., 2022 CASP checklist

## Data Availability

All data produced in the present work are contained in the manuscript

https://osf.io/6wf25/?view_only=6e1348facda14ac68e36a0c34bebe8b9

## Acknowledgements

This project was completed with support from Polygeia, a student-led global health think tank and Concentric Policies, a charity working to tackle non-communicable disease.

We acknowledge the support of Polygeia Leeds Branch Presidents Katherine Poon and Alex Abouharb, and Polygeia Executive Director Sanjush Dalmia. We also acknowledge Sam Emerman and J.T. Stanley from Concentric policies for providing the research question and objective, around which we built the project.

## Appendix 1

**Appendix 1:**
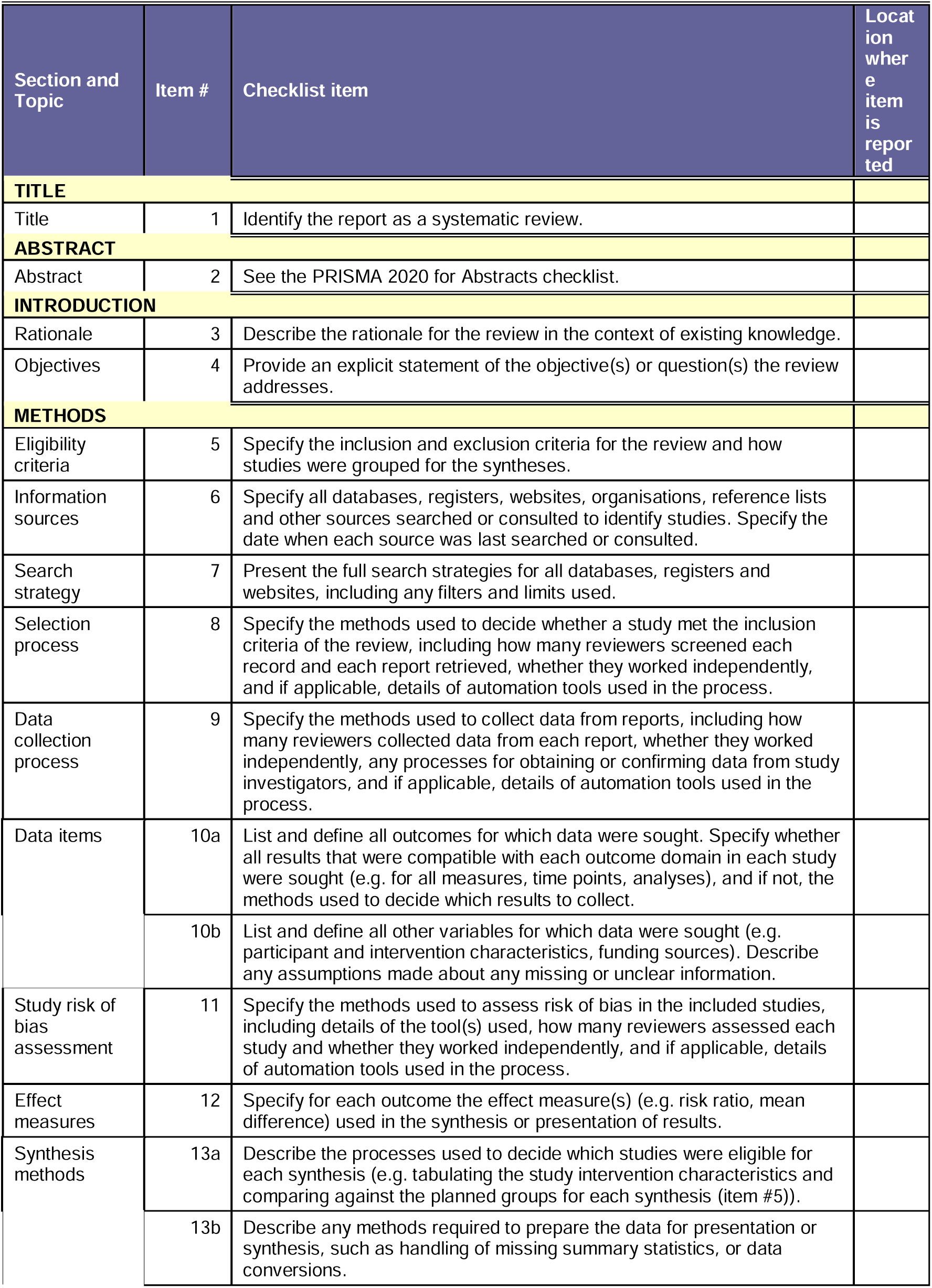
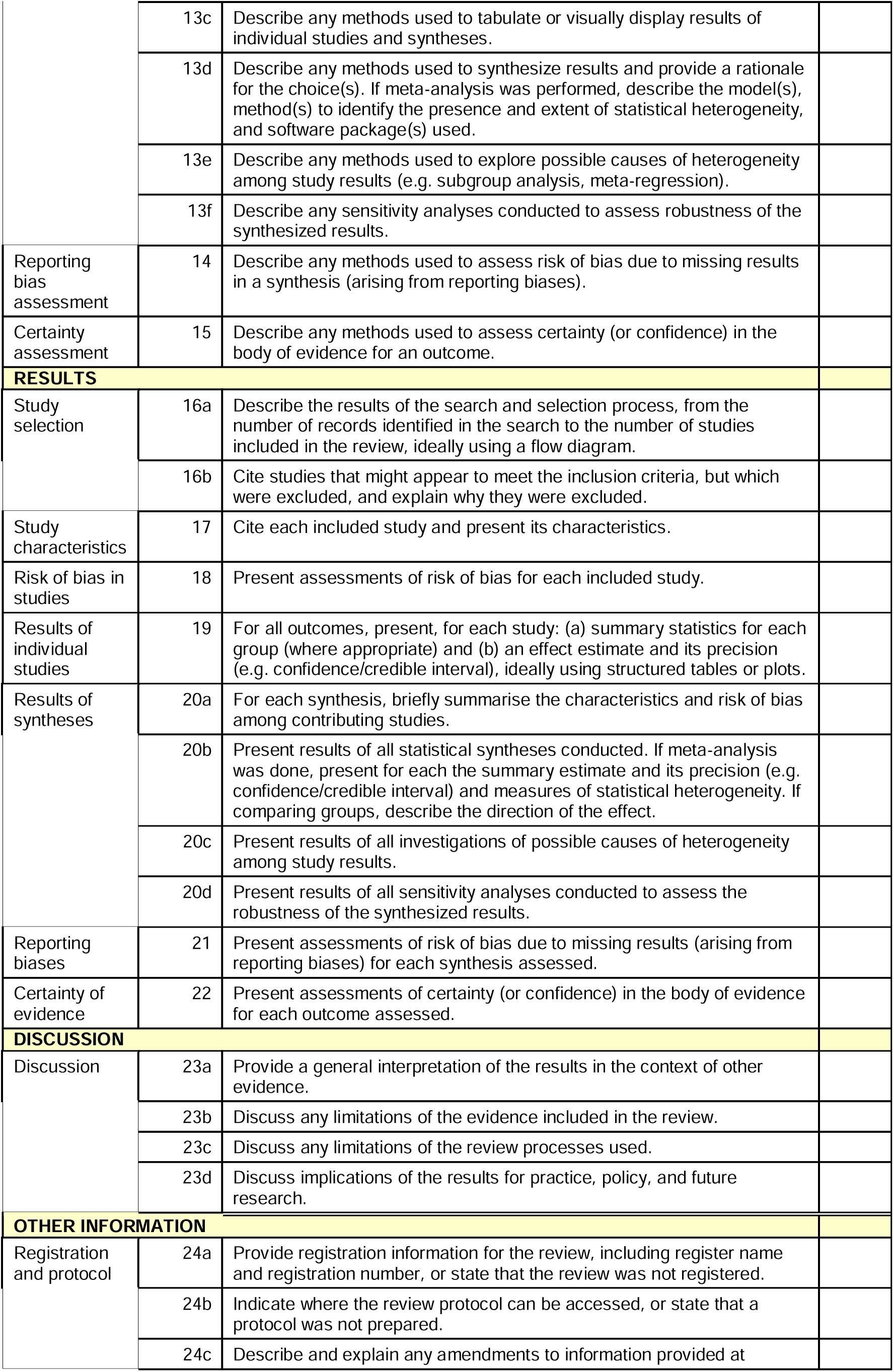
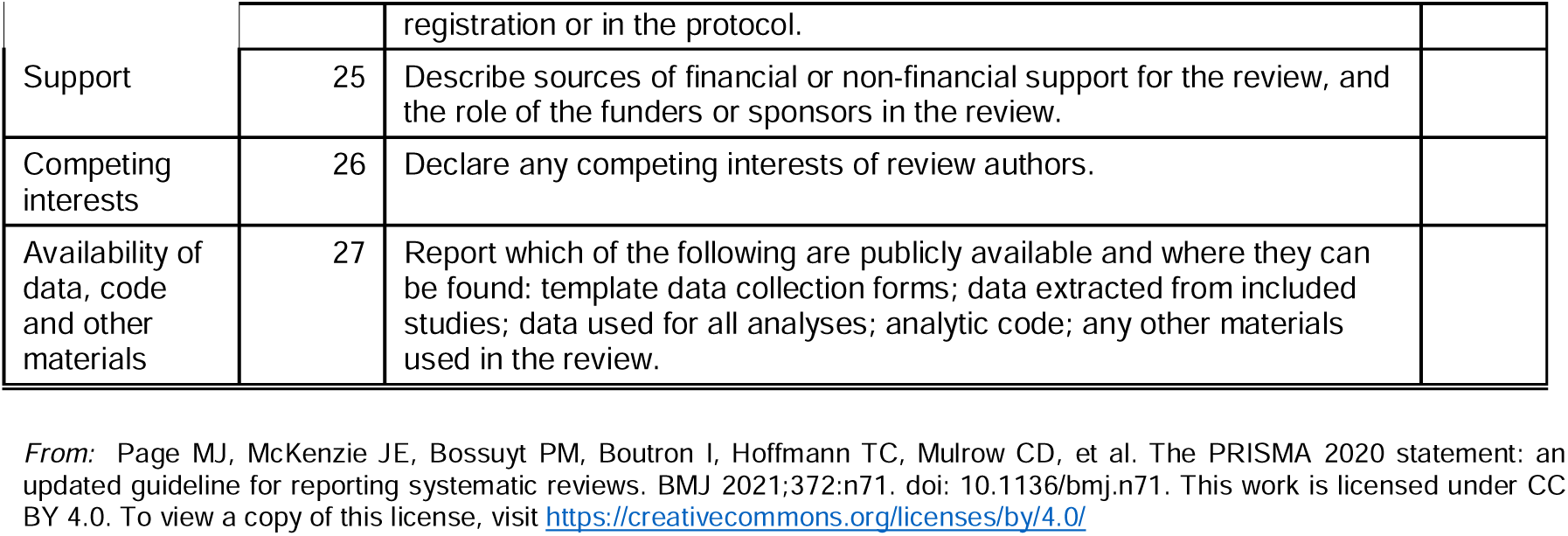
PRISMA 2020 Checklist.

| Section and Topic | Item # | Checklist item | Location where item is reported |
| --- | --- | --- | --- |
| **TITLE** |  |  |  |
| Title | 1 | Identify the report as a systematic review. |  |
| **ABSTRACT** |  |  |  |
| Abstract | 2 | See the PRISMA 2020 for Abstracts checklist. |  |
| **INTRODUCTION** |  |  |  |
| Rationale | 3 | Describe the rationale for the review in the context of existing knowledge. |  |
| Objectives | 4 | Provide an explicit statement of the objective(s) or question(s) the review addresses. |  |
| **METHODS** |  |  |  |
| Eligibility criteria | 5 | Specify the inclusion and exclusion criteria for the review and how studies were grouped for the syntheses. |  |
| Information sources | 6 | Specify all databases, registers, websites, organisations, reference lists and other sources searched or consulted to identify studies. Specify the date when each source was last searched or consulted. |  |
| Search strategy | 7 | Present the full search strategies for all databases, registers and websites, including any filters and limits used. |  |
| Selection process | 8 | Specify the methods used to decide whether a study met the inclusion criteria of the review, including how many reviewers screened each record and each report retrieved, whether they worked independently, and if applicable, details of automation tools used in the process. |  |
| Data collection process | 9 | Specify the methods used to collect data from reports, including how many reviewers collected data from each report, whether they worked independently, any processes for obtaining or confirming data from study investigators, and if applicable, details of automation tools used in the process. |  |
| Data items | 10a | List and define all outcomes for which data were sought. Specify whether all results that were compatible with each outcome domain in each study were sought (e.g. for all measures, time points, analyses), and if not, the methods used to decide which results to collect. |  |
|  | 10b | List and define all other variables for which data were sought (e.g. participant and intervention characteristics, funding sources). Describe any assumptions made about any missing or unclear information. |  |
| Study risk of bias assessment | 11 | Specify the methods used to assess risk of bias in the included studies, including details of the tool(s) used, how many reviewers assessed each study and whether they worked independently, and if applicable, details of automation tools used in the process. |  |
| Effect measures | 12 | Specify for each outcome the effect measure(s) (e.g. risk ratio, mean difference) used in the synthesis or presentation of results. |  |
| Synthesis methods | 13a | Describe the processes used to decide which studies were eligible for each synthesis (e.g. tabulating the study intervention characteristics and comparing against the planned groups for each synthesis (item #5)). |  |
|  | 13b | Describe any methods required to prepare the data for presentation or synthesis, such as handling of missing summary statistics, or data conversions. |  |
|  | 13c | Describe any methods used to tabulate or visually display results of individual studies and syntheses. |  |
|  | 13d | Describe any methods used to synthesize results and provide a rationale for the choice(s). If meta-analysis was performed, describe the model(s), method(s) to identify the presence and extent of statistical heterogeneity, and software package(s) used. |  |
|  | 13e | Describe any methods used to explore possible causes of heterogeneity among study results (e.g. subgroup analysis, meta-regression). |  |
|  | 13f | Describe any sensitivity analyses conducted to assess robustness of the synthesized results. |  |
| Reporting bias assessment | 14 | Describe any methods used to assess risk of bias due to missing results in a synthesis (arising from reporting biases). |  |
| Certainty assessment | 15 | Describe any methods used to assess certainty (or confidence) in the body of evidence for an outcome. |  |
| **RESULTS** |  |  |  |
| Study selection | 16a | Describe the results of the search and selection process, from the number of records identified in the search to the number of studies included in the review, ideally using a flow diagram. |  |
|  | 16b | Cite studies that might appear to meet the inclusion criteria, but which were excluded, and explain why they were excluded. |  |
| Study characteristics | 17 | Cite each included study and present its characteristics. |  |
| Risk of bias in studies | 18 | Present assessments of risk of bias for each included study. |  |
| Results of individual studies | 19 | For all outcomes, present, for each study: (a) summary statistics for each group (where appropriate) and (b) an effect estimate and its precision (e.g. confidence/credible interval), ideally using structured tables or plots. |  |
| Results of syntheses | 20a | For each synthesis, briefly summarise the characteristics and risk of bias among contributing studies. |  |
|  | 20b | Present results of all statistical syntheses conducted. If meta-analysis was done, present for each the summary estimate and its precision (e.g. confidence/credible interval) and measures of statistical heterogeneity. If comparing groups, describe the direction of the effect. |  |
|  | 20c | Present results of all investigations of possible causes of heterogeneity among study results. |  |
|  | 20d | Present results of all sensitivity analyses conducted to assess the robustness of the synthesized results. |  |
| Reporting biases | 21 | Present assessments of risk of bias due to missing results (arising from reporting biases) for each synthesis assessed. |  |
| Certainty of evidence | 22 | Present assessments of certainty (or confidence) in the body of evidence for each outcome assessed. |  |
| **DISCUSSION** |  |  |  |
| Discussion | 23a | Provide a general interpretation of the results in the context of other evidence. |  |
|  | 23b | Discuss any limitations of the evidence included in the review. |  |
|  | 23c | Discuss any limitations of the review processes used. |  |
|  | 23d | Discuss implications of the results for practice, policy, and future research. |  |
| **OTHER INFORMATION** |  |  |  |
| Registration and protocol | 24a | Provide registration information for the review, including register name and registration number, or state that the review was not registered. |  |
|  | 24b | Indicate where the review protocol can be accessed, or state that a protocol was not prepared. |  |
|  | 24c | Describe and explain any amendments to information provided at registration or in the protocol. |  |
| Support | 25 | Describe sources of financial or non-financial support for the review, and the role of the funders or sponsors in the review. |  |
| Competing interests | 26 | Declare any competing interests of review authors. |  |
| Availability of data, code and other materials | 27 | Report which of the following are publicly available and where they can be found: template data collection forms; data extracted from included studies; data used for all analyses; analytic code; any other materials used in the review. |  |

*From:* Page MJ, McKenzie JE, Bossuyt PM, Boutron I, Hoffmann TC, Mulrow CD, et al. The PRISMA 2020 statement: an updated guideline for reporting systematic reviews. *BMJ* 2021;372:n71. doi: 10.1136/bmj.n71. This work is licensed under CC BY 4.0. To view a copy of this license, visit https://creativecommons.org/licenses/by/4.0/

## Appendix 2

**Appendix 2:** Databases and query strings used to conduct the search strategy in this review.

| Database | Date Searched | Query String |
| --- | --- | --- |
| Medline | 08/08/2024 | ("health personnel"[MeSH Terms] OR "delivery of health care"[MeSH Terms] OR HCW[Text Word] OR HCWs[Text Word] OR Health staff[Text Word] OR Healthcare staff[Text Word] OR Healthcare professionals[Text Word] OR Health workers[Text Word] OR Healthcare workers[Text Word]) AND ("africa south of the sahara"[MeSH Terms] OR Sub-Saharan Africa[Text Word] OR "resource-limited settings"[MeSH Terms] OR Low Resource Settings[Text Word] OR "developing countries"[MeSH Terms] OR (low middle income countries[Text Word] OR low middle income countries settings[Text Word] OR low middle income countries this[Text Word] OR low middle income country[Text Word] OR low middle income country hospital[Text Word] OR low middle income country setting[Text Word] OR low middle income country settings[Text Word])) AND ("smoking cessation"[MeSH Terms] OR "tobacco use cessation"[MeSH Terms] OR "tobacco use cessation devices"[MeSH Terms] OR "tobacco use"[MeSH Terms] OR "tobacco control"[MeSH Terms] OR smoking cessation[Text Word] OR tobacco use cessation[Text Word] OR smoking abstinence[Text Word] OR "smoking prevention"[MeSH Terms]) AND medline[sb] |
| PubMed | 09/08/2024 | ("health personnel"[MeSH Terms] OR "delivery of health care"[MeSH Terms] OR HCW[Text Word] OR HCWs[Text Word] OR Health staff[Text Word] OR Healthcare staff[Text Word] OR Healthcare professionals[Text Word] OR Health workers[Text Word] OR Healthcare workers[Text Word]) AND ("africa south of the sahara"[MeSH Terms] OR Sub-Saharan Africa[Text Word] OR "resource-limited settings"[MeSH Terms] OR Low Resource Settings[Text Word] OR "developing countries"[MeSH Terms] OR (low middle income countries[Text Word] OR low middle income countries settings[Text Word] OR low middle income countries this[Text Word] OR low middle income country[Text Word] OR low middle income country hospital[Text Word] OR low middle income country setting[Text Word] OR low middle income country settings[Text Word])) AND ("smoking cessation"[MeSH Terms] OR "tobacco use cessation"[MeSH Terms] OR "tobacco use cessation devices"[MeSH Terms] OR "tobacco use"[MeSH Terms] OR "tobacco control"[MeSH Terms] OR smoking cessation[Text Word] OR tobacco use cessation[Text Word] OR smoking abstinence[Text Word] OR "smoking prevention"[MeSH Terms]) |
| Cochrane Library | 04/08/2024 | (MeSH descriptor: [Health Personnel] OR (health* NEXT ("Staff" OR professional* OR Worker*)):ti,ab,kw OR (HCW*):ti,ab,kw) AND (MeSH descriptor: [Africa South of the Sahara] OR sub-Sahar* NEXT(Africa)):ti,ab,kw) AND (MeSH |
|  |  | descriptor: [Tobacco Use Cessation Devices] OR MeSH descriptor: [Tobacco Use Cessation] OR MeSH descriptor: [Tobacco Control] OR ("tobacco" AND (Quit* OR "Abstinence" OR prevent*)):ti,ab,kw |
| Ovid | 07/08/2024 | (Health* staff) OR (health* professional) OR (health* worker*) OR HCW AND (smoking AND (cessation OR abstinence OR quit* OR control)) OR (tobacco AND (cessation OR abstinence OR quit* OR control) AND (sub-Sahar* OR Africa) OR (low-income OR low-resource) |
| Scopus | 11/08/2024 | (TITLE-ABS-KEY("health personnel" OR "delivery of health care" OR HCW OR HCWs OR "Health staff" OR "Healthcare staff" OR "Healthcare professionals" OR "Health workers" OR "Healthcare workers")) AND (TITLE-ABS-KEY("africa south of the sahara" OR "Sub-Saharan Africa" OR "resource-limited settings" OR "Low Resource Settings" OR "developing countries" OR "low middle income countries" OR "low middle income country" OR "low middle income country hospital" OR "low middle income country setting" OR "low middle income country settings")) AND (TITLE-ABS-KEY("smoking cessation" OR "tobacco use cessation" OR "tobacco use cessation devices" OR "tobacco use" OR "tobacco control" OR "smoking cessation" OR "tobacco use cessation" OR "smoking abstinence" OR "smoking prevention")) |

