## Supplementary material for "Efficacy and Best Practices of Health-care worker Smoking Cessation Treatment in Sub-Saharan Africa": Louwagie et al., 2014 CASP checklist

Critical Appraisal Skills Programme

**CASP Randomised Controlled Trial Standard Checklist:**
11 questions to help you make sense of a randomised controlled trial (RCT)

**Main issues for consideration:** Several aspects need to be considered when appraising a randomised controlled trial:

- Is the basic study design valid for a randomised controlled trial? (Section A)
- Was the study methodologically sound? (Section B)
- What are the results? (Section C)
- Will the results help locally? (Section D)

The 11 questions in the checklist are designed to help you think about these aspects systematically.

**How to use this appraisal tool:** The first three questions (Section A) are screening questions about the validity of the basic study design and can be answered quickly. If, in light of your responses to Section A, you think the study design is valid, continue to Section B to assess whether the study was methodologically sound and if it is worth continuing with the appraisal by answering the remaining questions in Sections C and D.

Record 'Yes', 'No' or 'Can't tell' in response to the questions. Prompts below all but one of the questions highlight the issues it is important to consider. Record the reasons for your answers in the space provided. As CASP checklists were designed to be used as educational/teaching tools in a workshop setting, we do not recommend using a scoring system.

**About CASP Checklists:** The CASP RCT checklist was originally based on JAMA Users' guides to the medical literature 1994 (adapted from Guyatt GH, Sackett DL and Cook DJ), and piloted with healthcare practitioners. This version has been updated taking into account the CONSORT 2010 guideline (http://www.consort-statement.org/consort-2010, accessed 16 September 2020).

**Citation:** CASP recommends using the Harvard style, i.e., *Critical Appraisal Skills Programme (2021). CASP (insert name of checklist i.e. Randomised Controlled Trial) Checklist. [online] Available at: insert URL. Accessed: insert date accessed.*

©CASP this work is licensed under the Creative Commons Attribution – Non-Commercial- Share A like. To view a copy of this licence, visit https://creativecommons.org/licenses/by-sa/4.0/

CASP
Critical Appraisal
Skills Programme

**Study and citation:** Efficacy of brief motivational interviewing on smoking cessation at tuberculosis clinics in Tshwane, South Africa: a randomized controlled trial

[Efficacy of brief motivational interviewing on smoking cessation at tuberculosis clinics in Tshwane, South Africa: a randomized controlled trial - PubMed (nih.gov)]

(Louwagie et al., 2014)

Louwagie, G.M.C., Okuyemi, K.S. and Ayo-Yusuf, O.A. 2014. Efficacy of brief motivational interviewing on smoking cessation at tuberculosis clinics in Tshwane, South Africa: a randomized controlled trial. *Addiction*. **109**(11), pp.1942–1952.

### Section A: Is the basic study design valid for a randomised controlled trial?

| | Question | Yes | No | Can't tell |
|---|---|---|---|---|
| **1.** | Did the study address a clearly focused research question?<br>*CONSIDER:*<br>• *Was the study designed to assess the outcomes of an intervention?*<br>• *Is the research question 'focused' in terms of:*<br> • *Population studied*<br> • *Intervention given*<br> • *Comparator chosen*<br> • *Outcomes measured?* | ☒ | ☐ | ☐ |
| 2. | Was the assignment of participants to interventions randomised?<br>*CONSIDER:*<br>• *How was randomisation carried out? Was the method appropriate?*<br>• *Was randomisation sufficient to eliminate systematic bias?*<br>• *Was the allocation sequence concealed from investigators and participants?* | ☒ | ☐ | ☐ |
| **3.** | Were all participants who entered the study accounted for at its conclusion?<br>*CONSIDER:*<br>• *Were losses to follow-up and exclusions after randomisation accounted for?*<br>• *Were participants analysed in the study groups to which they were randomised (intention-to-treat analysis)?*<br>• *Was the study stopped early? If so, what was the reason?* | ☒ | ☐ | ☐ |

### Section B: Was the study methodologically sound?

| | Question | Yes | No | Can't tell |
|---|---|---|---|---|
| 4. | • Were the participants 'blind' to intervention they were given? | ☐ | ☒ | ☐ |
| | • Were the investigators 'blind' to the intervention they were giving to participants? | ☒ | ☐ | ☐ |
| | • Were the people assessing/analysing outcome/s 'blinded'? | ☒ | ☐ | ☐ |

| | |
|---|---|
| 5. Were the study groups similar at the start of the randomised controlled trial?<br>*CONSIDER:*<br>• *Were the baseline characteristics of each study group (e.g. age, sex, socio-economic group) clearly set out?*<br>• *Were there any differences between the study groups that could affect the outcome/s?* | Yes ☒ No ☐ Can't tell ☐ |

Critical Appraisal Skills Programme

| | | | |
|---|---|---|---|
| **6.** Apart from the experimental intervention, did each study group receive the same level of care (that is, were they treated equally)?<br><br>*CONSIDER:*<br>• *Was there a clearly defined study protocol?*<br>• *If any additional interventions were given (e.g. tests or treatments), were they similar between the study groups?*<br>• *Were the follow-up intervals the same for each study group?* | Yes ☒ | No ☐ | Can't tell ☐ |

Section C: What are the results?

| | | | |
|---|---|---|---|
| 7. Were the effects of intervention reported comprehensively?<br><br>*CONSIDER:*<br>• *Was a power calculation undertaken?*<br>• *What outcomes were measured, and were they clearly specified?*<br>• *How were the results expressed? For binary outcomes, were relative and absolute effects reported?*<br>• *Were the results reported for each outcome in each study group at each follow-up interval?*<br>• *Was there any missing or incomplete data?*<br>• *Was there differential drop-out between the study groups that could affect the results?*<br>• *Were potential sources of bias identified?*<br>• *Which statistical tests were used?*<br>• *Were p values reported?* | Yes ☒ | No ☐ | Can't tell ☐ |
| 8. Was the precision of the estimate of the intervention or treatment effect reported?<br><br>*CONSIDER:*<br>• *Were confidence intervals (CIs) reported?* | Yes ☒ | No ☐ | Can't tell ☐ |
| 9. Do the benefits of the experimental intervention outweigh the harms and costs?<br><br>*CONSIDER:*<br>• *What was the size of the intervention or treatment effect?*<br>• *Were harms or unintended effects reported for each study group?*<br>• *Was a cost-effectiveness analysis undertaken? (Cost-effectiveness analysis allows a comparison to be made between different interventions used in the care of the same condition or problem.)* | Yes ☐ | No ☐ | Can't tell ☒ |

Critical Appraisal Skills Programme

| Section D: Will the results help locally? |
| --- |

| | | | | |
| --- | --- | --- | --- | --- |
| 10. | Can the results be applied to your local population/in your context?<br><br>*CONSIDER:*<br>• *Are the study participants similar to the people in your care?*<br>• *Would any differences between your population and the study participants alter the outcomes reported in the study?*<br>• *Are the outcomes important to your population?*<br>• *Are there any outcomes you would have wanted information on that have not been studied or reported?*<br>• *Are there any limitations of the study that would affect your decision?* | Yes ☒ | No ☐ | Can't tell ☐ |
| 11. | Would the experimental intervention provide greater value to the people in your care than any of the existing interventions?<br><br>*CONSIDER:*<br>• *What resources are needed to introduce this intervention taking into account time, finances, and skills development or training needs?*<br>• *Are you able to disinvest resources in one or more existing interventions in order to be able to re-invest in the new intervention?* | Yes ☒ | No ☐ | Can't tell ☐ |

APPRAISAL SUMMARY: *Record key points from your critical appraisal in this box. What is your conclusion about the paper? Would you use it to change your practice or to recommend changes to care/interventions used by your organisation? Could you judiciously implement this intervention without delay?*

Strengths of the Study:

- Using Randomisation by allocating the patient in intervention and control group through envelope concealment.
- Addressed a prevalent issue in low resource setting (HIV, TB and smoking).
- The use of Lay Healthcare workers (LHCWs) for brief motivational interviewing (MI) is a cheap strategy to implement and has potential to be scaled in low resource settings.
- Statistically significant results showing higher smoking cessation rates in the intervention group.

Limitations of the Study:

- Lack of blinding for participants and LHCWs, which introduces potential bias.
- High dropout rates – reduces the accuracy of the results.
- Single-session MI and absence of pharmacotherapy. This may limit the intervention's long-term effectiveness and how it can be supplemented by other strategies.
- Short follow-up period of six months doesn't assess the long-term relapse of smoking.

Conclusion:

- The study shows that brief MI by LHCWs can be effective in helping TB patients quit smoking, particularly in resource-limited settings, where other interventions like pharmacotherapy might be too costly.
- I would consider using this strategy in low-resource settings, but I would also look for research comparing its effectiveness and costs with other strategies.
- This intervention could be easily implemented as it is not too costly.
