## Supplementary material for "Efficacy and Best Practices of Health-care worker Smoking Cessation Treatment in Sub-Saharan Africa": Tamirat, 2021 CASP checklist

CASP
Critical Appraisal Skills Programme

www.casp-uk.net

Summertown Pavilion, Middle Way Oxford OX2 7LG

### CASP Checklist: 12 questions to help you make sense of a Cohort Study

**How to use this appraisal tool:** Three broad issues need to be considered when appraising a cohort study:

- Are the results of the study valid? (Section A)
- What are the results? (Section B)
- Will the results help locally? (Section C)

The 12 questions on the following pages are designed to help you think about these issues systematically. The first two questions are screening questions and can be answered quickly. If the answer to both is "yes", it is worth proceeding with the remaining questions. There is some degree of overlap between the questions, you are asked to record a "yes", "no" or "can't tell" to most of the questions. A number of italicised prompts are given after each question. These are designed to remind you why the question is important. Record your reasons for your answers in the spaces provided.

**About:** These checklists were designed to be used as educational pedagogic tools, as part of a workshop setting, therefore we do not suggest a scoring system. The core CASP checklists (randomised controlled trial & systematic review) were based on JAMA 'Users' guides to the medical literature 1994 (adapted from Guyatt GH, Sackett DL, and Cook DJ), and piloted with health care practitioners.

For each new checklist, a group of experts were assembled to develop and pilot the checklist and the workshop format with which it would be used. Over the years overall adjustments have been made to the format, but a recent survey of checklist users reiterated that the basic format continues to be useful and appropriate.

**Referencing:** we recommend using the Harvard style citation, i.e.: *Critical Appraisal Skills Programme (2018). CASP (insert name of checklist i.e. Cohort Study) Checklist. [online] Available at: URL. Accessed: Date Accessed.*

©CASP this work is licensed under the Creative Commons Attribution – Non-Commercial-Share A like. To view a copy of this license, visit http://creativecommons.org/licenses/by-nc-sa/3.0/ www.casp-uk.net

Critical Appraisal Skills Programme

Paper for appraisal and reference: Health Workers' Practice Towards Smoking Cessation Ir

#### Section A: Are the results of the study valid?

1. Did the study address a clearly focused issue?

Yes ✔ | Can't Tell ☐ | No ☐

HINT: A question can be 'focused' in terms of
- the population studied
- the risk factors studied
- is it clear whether the study tried to detect a beneficial or harmful effect
- the outcomes considered

Comments: Study focused on Healthcare Worker's practice of using the 5A's model as an intervention for smoking cessation. It was carried out in the Hadiya zone, Southern Ethiopia. It examined variables influencing smoking cessation, including gender, training, knowledge, and years of service

2. Was the cohort recruited in an acceptable way?

Yes ☐ | Can't Tell ✔ | No ☐

HINT: Look for selection bias which might compromise the generalisability of the findings:
- was the cohort representative of a defined population
- was there something special about the cohort
- was everybody included who should have been

Comments: Consisted of healthcare workers from 4 different hospitals in the Hadiya zone using a random sampling technique with proportional representation from each hospital.
Difficult to determine if the conclusion will be the same if the study was conducted in other countries and in populations where resources aren't limited.

Is it worth continuing?

3. Was the exposure accurately measured to minimise bias?

| | |
|---|---|
| Yes | ✔ |
| Can't Tell | |
| No | |

HINT: Look for measurement or classification bias:

- did they use subjective or objective measurements
- do the measurements truly reflect what you want them to (have they been validated)
- were all the subjects classified into exposure groups using the same procedure

Comments: The exposure in this study was the health workers' smoking cessation practices, which were measured using a self-administered questionnaire centred around the 5A's model.
Training was also provided for the data collectors, and the questionnaire was tested for clarity to improve accuracy.

4. Was the outcome accurately measured to minimise bias?

| | |
|---|---|
| Yes | ✔ |
| Can't Tell | |
| No | |

HINT: Look for measurement or classification bias:

- did they use subjective or objective measurements
- do the measurements truly reflect what you want them to (have they been validated)
- has a reliable system been established for detecting all the cases (for measuring disease occurrence)
- were the measurement methods similar in the different groups
- were the subjects and/or the outcome assessor blinded to exposure (does this matter)

Comments: Data collection through the questionnaire was standardised by training the nurses to ensure consistency across all four hospitals. Therefore reducing the risk of measurement bias.

5. (a) Have the authors identified all important confounding factors?

Yes ☑
Can't Tell ☐
No ☐

HINT:
- list the ones you think might be important, and ones the author missed

Comments: Key confounding factors identified by the author were gender, knowledge, training, and years of service among healthcare workers. These are all factors directly linked to smoking cessation practices.

5. (b) Have they taken account of the confounding factors in the design and/or analysis?

Yes ☑
Can't Tell ☐
No ☐

HINT:
- look for restriction in design, and techniques e.g. modelling, stratified-, regression-, or sensitivity analysis to correct, control or adjust for confounding factors

Comments: Multivariable binary logistic regression was used to adjust for confounding variables such as training, knowledge, gender, and years of service. This ensured a more accurate analysis.

6. (a) Was the follow up of subjects complete enough?

Yes ☑
Can't Tell ☐
No ☐

HINT: Consider
- the good or bad effects should have had long enough to reveal themselves
- the persons that are lost to follow-up may have different outcomes than those available for assessment
- in an open or dynamic cohort, was there anything special about the outcome of the people leaving, or the exposure of the people entering the cohort

6. (b) Was the follow up of subjects long enough?

Yes ☑
Can't Tell ☐
No ☐

Comments: The response rate was 97.2%, which is sufficient for a cross-sectional design.
The study's data collection period lasted for two months, which provided enough time for the questionnaire to be completed.

#### Section B: What are the results?

7. What are the results of this study?

HINT: Consider
- what are the bottom line results
- have they reported the rate or the proportion between the exposed/unexposed, the ratio/rate difference
- how strong is the association between exposure and outcome (RR)
- what is the absolute risk reduction (ARR)

Comments: 97% of healthcare workers were identified as having poor smoking cessation practices, leaving only 3% with good practice.
The study found that being male, trained, having good knowledge, and fewer than 10 years of service were associated with better smoking cessation practices.

8. How precise are the results?

HINT:
- look for the range of the confidence intervals, if given

Comments: The results were significant for the factors listed below when analysed using logistic regression, indicating the precision of the associations.
The following associations were observed:
Male healthcare workers were 2.25 times more likely to practise smoking cessation interventions compared to females (AOR = 2.25, 95% CI 1.31–6.23).
Healthcare workers with formal training were 6.5 times more likely to engage in smoking cessation practices than those without training (AOR =

9. Do you believe the results?

- Yes ✔
- Can't Tell ☐
- No ☐

HINT: Consider
- big effect is hard to ignore
- can it be due to bias, chance or confounding
- are the design and methods of this study sufficiently flawed to make the results unreliable
- Bradford Hills criteria (e.g. time sequence, dose-response gradient, biological plausibility, consistency)

Comments: The results seem logical given the detailed methodology, high response rate, and the use of analysis tools to adjust for confounding factors. Training was also provided to the data collectors, contributing to the precision of the data collection process.

#### Section C: Will the results help locally?

10. Can the results be applied to the local population?

- Yes ✔
- Can't Tell ☐
- No ☐

HINT: Consider whether
- a cohort study was the appropriate method to answer this question
- the subjects covered in this study could be sufficiently different from your population to cause concern
- your local setting is likely to differ much from that of the study
- you can quantify the local benefits and harms

Comments: The results will most likely apply to similar low-resource settings in Sub-Saharan Africa.

11. Do the results of this study fit with other available evidence?

- Yes ☐
- Can't Tell ☐
- No ✔

Comments: For some aspects of the 5A's model, similar results were found in studies conducted globally, particularly in low- and middle-income countries like Kenya and India. However, the results differed in other aspects of the 5A's model in places like Catalonia and Italy. Overall, the findings present a [illegible]

12. What are the implications of this study for practice?

| | |
|---|---|
| Yes | ☐ |
| Can't Tell | ☑ |
| No | ☐ |

HINT: Consider
- one observational study rarely provides sufficiently robust evidence to recommend changes to clinical practice or within health policy decision making
- for certain questions, observational studies provide the only evidence
- recommendations from observational studies are always stronger when supported by other evidence

Comments: The study highlights the need to improve the practice of the 5A's model, as it is currently poorly practised. This may affect the policies and guidelines used in the Hadiya region and similar settings.
The usefulness of the 5A's model also needs to be robustly studied before implementation to research its effectiveness in smoking cessation.
