## Supplementary material for "Efficacy and Best Practices of Health-care worker Smoking Cessation Treatment in Sub-Saharan Africa": Rossouw and Filby, 2022 CASP checklist

Paper for appraisal and reference: Inequalities in successful tobacco cessation and tobacco c

**Section A: Are the results of the study valid?**

1. Did the study address a clearly focused issue?

| | |
|---|---|
| Yes | ✔ |
| Can't Tell | |
| No | |

HINT: A question can be 'focused' in terms of
- the population studied
- the risk factors studied
- is it clear whether the study tried to detect a beneficial or harmful effect
- the outcomes considered

Comments:

2. Was the cohort recruited in an acceptable way?

| | |
|---|---|
| Yes | ✔ |
| Can't Tell | |
| No | |

HINT: Look for selection bias which might compromise the generalisability of the findings:
- was the cohort representative of a defined population
- was there something special about the cohort
- was everybody included who should have been

Comments: Global Adult Tobacco Survey (GATS), which is designed to be nationally representative. Stratified Multi-Stage Cluster Sampling, Use of Country-Specific Weight

**Is it worth continuing?**

CASP
Critical Appraisal
Skills Programme

3. Was the exposure accurately measured to minimise bias?

Yes ☑ ✔
Can't Tell ☐
No ☐

HINT: Look for measurement or classification bias:
- did they use subjective or objective measurements
- do the measurements truly reflect what you want them to (have they been validated)
- were all the subjects classified into exposure groups using the same procedure

Comments:

4. Was the outcome accurately measured to minimise bias?

Yes ☑ ✔
Can't Tell ☐
No ☐

HINT: Look for measurement or classification bias:
- did they use subjective or objective measurements
- do the measurements truly reflect what you want them to (have they been validated)
- has a reliable system been established for detecting all the cases (for measuring disease occurrence)
- were the measurement methods similar in the different groups
- were the subjects and/or the outcome assessor blinded to exposure (does this matter)

Comments: Self-Reported Data,

5. (a) Have the authors identified all important confounding factors?

Yes ✔
Can't Tell ☐
No ☐

HINT:
- list the ones you think might be important, and ones the author missed

Comments: identified and controlled for many of the most important confounding factors that could influence tobacco cessation outcomes, such as wealth, education, demographic factors (age, gender, marital status), employment status, and knowledge about tobacco harms

5. (b) Have they taken account of the confounding factors in the design and/or analysis?

Yes ✔
Can't Tell ☐
No ☐

HINT:
- look for restriction in design, and techniques e.g. modelling, stratified-, regression-, or sensitivity analysis to correct, control or adjust for confounding factors

Comments:

6. (a) Was the follow up of subjects complete enough?

Yes ☐
Can't Tell ✔
No ☐

HINT: Consider
- the good or bad effects should have had long enough to reveal themselves
- the persons that are lost to follow-up may have different outcomes than those available for assessment
- in an open or dynamic cohort, was there anything special about the outcome of the people leaving, or the exposure of the people entering the cohort

6. (b) Was the follow up of subjects long enough?

Yes ☐
Can't Tell ✔
No ☐

Comments:

Section B: What are the results?

7. What are the results of this study?

HINT: Consider
- what are the bottom line results
- have they reported the rate or the proportion between the exposed/unexposed, the ratio/rate difference
- how strong is the association between exposure and outcome (RR)
- what is the absolute risk reduction (ARR)

Comments: Wealthier individuals in Cameroon, Ethiopia, Kenya, and Uganda are more likely to attempt quitting range from 0.100 in Kenya to 0.382 in Ethiopia on the CIE, all statistically significant.) Higher education levels are associated with more quit attempts, with significant disparities in countries like Ethiopia (0.222), Kenya (0.331), and Nigeria (0.151).
In Former Tobacco Users (TCF), Education also drives disparities

8. How precise are the results?

HINT:
- look for the range of the confidence intervals, if given

Comments: precise - highly significant p-values (most < 0.05 or 0.01), The standard errors in the study range from 0.024 to 0.065,

CASP
Critical Appraisal
Skills Programme

9. Do you believe the results?

Yes ✔
Can't Tell ☐
No ☐

HINT: Consider
- big effect is hard to ignore
- can it be due to bias, chance or confounding
- are the design and methods of this study sufficiently flawed to make the results unreliable
- Bradford Hills criteria (e.g. time sequence, dose-response gradient, biological plausibility, consistency)

Comments:

Section C: Will the results help locally?

10. Can the results be applied to the local population?

Yes ✔
Can't Tell ☐
No ☐

HINT: Consider whether
- a cohort study was the appropriate method to answer this question
- the subjects covered in this study could be sufficiently different from your population to cause concern
- your local setting is likely to differ much from that of the study
- you can quantify the local benefits and harms

Comments:

11. Do the results of this study fit with other available evidence?

Yes ✔
Can't Tell ☐
No ☐

Comments:

12. What are the implications of this study for practice?

Yes ✔

Can't Tell ☐

No ☐

HINT: Consider
- one observational study rarely provides sufficiently robust evidence to recommend changes to clinical practice or within health policy decision making
- for certain questions, observational studies provide the only evidence
- recommendations from observational studies are always stronger when supported by other evidence

Comments: less economically developed areas should be targeted for smoking cessation
