## Supplementary material for "Efficacy and Best Practices of Health-care worker Smoking Cessation Treatment in Sub-Saharan Africa": Du Plooy et al., 2016 CASP checklist

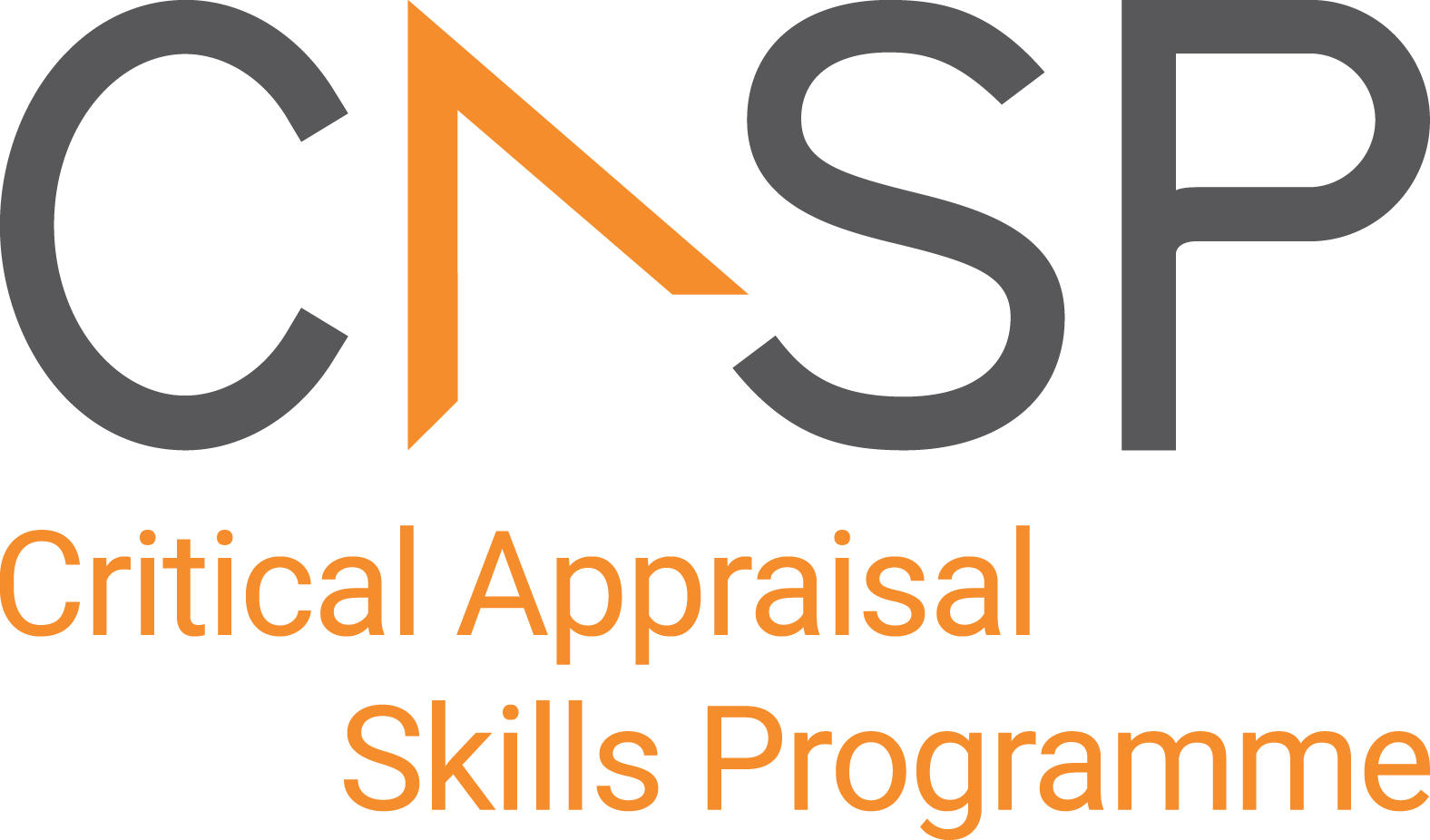
CASP Checklist:

For Cohort Studies

| **Paper Title:** | Cigarette smoking, nicotine dependence, and motivation to quit smoking in South African male psychiatric inpatients |
| --- | --- |
| **Author:** | Du Plooy et al., 2016 |
| **Web Link:** | https://bmcpsychiatry.biomedcentral.com/articles/10.1186/s12888-016-1123-z |

During critical appraisal, never make assumptions about what the researchers have done. If it is not possible to tell, use the “Can’t tell” response box. If you can’t tell, at best it means the researchers have not been explicit or transparent, but at worst it could mean the researchers have not undertaken a particular task or process. Once you’ve finished the critical appraisal, if there are a large number of “Can’t tell” responses, consider whether the findings of the study are trustworthy and interpret the results with caution.

| **Section A: Are the results valid?** | | |
| --- | --- | --- |
| 1. Did the study address a clearly focused issue? | ☐Yes ☐ No ☐ Can’t Tell | |
| *CONSIDER:*  *A question can be ‘focused’ in terms of*   - *the population studied* - *the risk factors studied* - *is it clear whether the study tried to detect a beneficial or harmful effect* - *the outcomes considered* | | |
| 2. Was the cohort recruited in an acceptable way? | ☐Yes ☐ No ☐ Can’t Tell | |
| *CONSIDER:*   - *Look for selection bias which might compromise the generalisability of the findings:* - *was the cohort representative of a defined population* - *was there something special about the cohort* - *was everybody included who should have been* | | |
| 3. Was the exposure accurately measured to minimise bias? | ☐Yes ☐ No ☐ Can’t Tell | |
| *CONSIDER:*  *Look for measurement or classification bias:*   - *did they use subjective or objective measurements* - *do the measurements truly reflect what you want them to (have they been validated)* - *were all the subjects classified into exposure groups using the same procedure* | | |
| 4. Was the outcome accurately measured to minimise bias? | ☐Yes ☐ No ☐ Can’t Tell | |
| *CONSIDER:*  *Look for measurement or classification bias:*   - *did they use subjective or objective measurements* - *do the measurements truly reflect what you want them to (have they been validated)* - *has a reliable system been established for detecting all the cases (for measuring disease occurrence)* - *were the measurement methods similar in the different groups* - *were the subjects and/or the outcome assessor blinded to exposure (does this matter)* | | |
| 5. (a) Have the authors identified all important confounding factors? | ☐Yes ☐ No ☐ Can’t Tell | |
| *CONSIDER:*   - *list the ones you think might be important, and ones the author missed* | | |
| b) Have they taken account of the confounding factors in the design and/or analysis? | ☐Yes ☐ No ☐ Can’t Tell | |
| *CONSIDER:*   - *look for restriction in design, and techniques e.g. modelling, stratified-, regression-, or sensitivity analysis to correct, control or adjust for confounding factors* | | |
| 6. a) Was the follow up of subjects complete enough? | | ☐Yes ☐ No ☐ Can’t Tell |
| *CONSIDER:*   - *the persons that are lost to follow-up may have different outcomes than those available for assessment* - *in an open or dynamic cohort, was there anything special about the outcome of the people leaving, or the exposure of the people entering the cohort* | | |
| b) Was the follow up of subjects long enough? | | ☐Yes ☐ No ☐ Can’t Tell |
| *CONSIDER:*   - *the good or bad effects should have had long enough to reveal themselves* | | |
| **Section B: What are the results?** | | |
| 7. What are the results of this study? | ☐Yes ☐ No ☐ Can’t Tell | |
| *CONSIDER:*   - *what are the bottom line results* - *have they reported the rate or the proportion between the exposed/unexposed, the ratio/rate difference* - *how strong is the association between exposure and outcome (RR)* - *what is the absolute risk reduction (ARR)* | | |
| 8. How precise are the results? | ☐Yes ☐ No ☐ Can’t Tell | |
| *CONSIDER:*   - *look for the range of the confidence intervals, if given* | | |
| 9. Do you believe the results? | ☐Yes ☐ No ☐ Can’t Tell | |
| *CONSIDER:*   - *big effect is hard to ignore* - *can it be due to bias, chance or confounding* - *are the design and methods of this study sufficiently flawed to make the results unreliable* - *Bradford Hills criteria (e.g. time sequence, dose-response gradient, biological plausibility, consistency)* | | |
| **Section C: Will the results help locally?** | | |
| 10. Can the results be applied to the local population? | ☐Yes ☐ No ☐ Can’t Tell | |
| *CONSIDER:*   - *Is a cohort study the appropriate method to answer this question* - *If the subjects covered in this study could be sufficiently different from your population to cause concern* - *If your local setting is likely to differ much from that of the study* - *If you can quantify the local benefits and harms* | | |
| 11. Do the results of this study fit with other available evidence? | | ☐Yes ☐ No ☐ Can’t Tell |
| 12. What are the implications of this study for practice? | | ☐Yes ☐ No ☐ Can’t Tell |
| *CONSIDER:*   - *one observational study rarely provides sufficiently robust evidence to recommend changes to clinical practice or within health policy decision making* - *for certain questions, observational studies provide the only evidence* - *recommendations from observational studies are always stronger when supported by other evidence* | | |

| **APPRAISAL SUMMARY**: *List key points from your critical appraisal that need to be considered when assessing the validity of the results and their usefulness in decision-making.* | | |
| --- | --- | --- |
| **Positive/Methodologically sound** | **Negative/Relatively poor methodology** | **Unknowns** |
| The study is methodologically sound in its design and use of validated tools. | Lack of comprehensive consideration of all possible confounders and a lack of female representation. | Long-term follow-up findings in the cohort |

**Referencing recommendation:**

CASP recommends using the Harvard style referencing, which is an author/date method. Sources are cited within the body of your assignment by giving the name of the author(s) followed by the date of publication. All other details about the publication are given in the list of references or bibliography at the end.

Example:

*Critical Appraisal Skills Programme (2024). CASP (insert name of checklist i.e. qualitative studies Checklist.) [online] Available at: insert URL. Accessed: insert date accessed.*

**Creative Commons**

©CASP this work is licensed under the Creative Commons Attribution – Non-Commercial- Share A like. To view a copy of this licence, visit <https://creativecommons.org/licenses/by-nc-sa/4.0/>

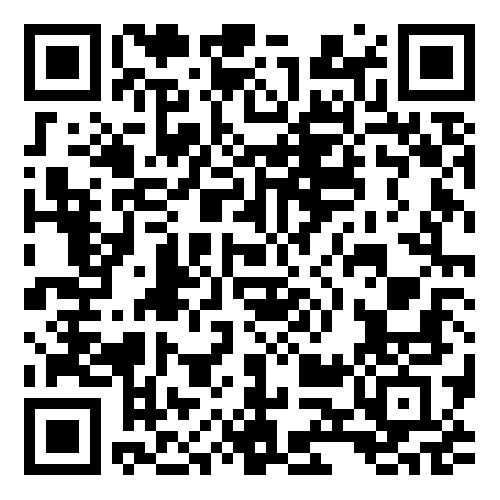
**Need further training on evidence-based decision making?** Our online training courses are helpful for healthcare educational researchers and any other learners who:

- Need to critically appraise and stay abreast of the healthcare research literature as part of their clinical duties.
- Are considering carrying out research & developing their own research projects.
- Make decisions in their role, whether that be policy making or patient facing.

**Benefits of CASP Training:**

- Affordable – courses start from as little as £6
- Professional training – leading experts in critical appraisal training
- Self-directed study – complete each course in your own time
- 12 months access – revisit areas you aren’t sure of and revise
- CPD certification - after each completed module

Scan the QR code below or visit <https://casp-uk.net/critical-appraisal-online-training-courses/> for more information and to start learning more.
