## Supplementary material for "Efficacy and Best Practices of Health-care worker Smoking Cessation Treatment in Sub-Saharan Africa": Mahoto et al., 2023 CASP checklist

CASP
Critical Appraisal Skills Programme

www.casp-uk.net

Summertown Pavilion, Middle Way Oxford OX2 7LG

### CASP Checklist: 10 questions to help you make sense of a Qualitative research

**How to use this appraisal tool:** Three broad issues need to be considered when appraising a qualitative study:

- Are the results of the study valid? (Section A)
- What are the results? (Section B)
- Will the results help locally? (Section C)

The 10 questions on the following pages are designed to help you think about these issues systematically. The first two questions are screening questions and can be answered quickly. If the answer to both is “yes”, it is worth proceeding with the remaining questions. There is some degree of overlap between the questions, you are asked to record a “yes”, “no” or “can’t tell” to most of the questions. A number of italicised prompts are given after each question. These are designed to remind you why the question is important. Record your reasons for your answers in the spaces provided.

**About:** These checklists were designed to be used as educational pedagogic tools, as part of a workshop setting, therefore we do not suggest a scoring system. The core CASP checklists (randomised controlled trial & systematic review) were based on JAMA 'Users’ guides to the medical literature 1994 (adapted from Guyatt GH, Sackett DL, and Cook DJ), and piloted with health care practitioners.

For each new checklist, a group of experts were assembled to develop and pilot the checklist and the workshop format with which it would be used. Over the years overall adjustments have been made to the format, but a recent survey of checklist users reiterated that the basic format continues to be useful and appropriate.

**Referencing:** we recommend using the Harvard style citation, i.e.: *Critical Appraisal Skills Programme (2018). CASP (insert name of checklist i.e. Qualitative) Checklist. [online] Available at: URL. Accessed: Date Accessed.*

©CASP this work is licensed under the Creative Commons Attribution – Non-Commercial-Share A like. To view a copy of this license, visit http://creativecommons.org/licenses/by-nc-sa/3.0/ www.casp-uk.net

CASP
Critical Appraisal
Skills Programme

Paper for appraisal and reference: Barriers to the provision of smoking cessation intervention/[illegible]

#### Section A: Are the results valid?

1. Was there a clear statement of the aims of the research?

| | |
|---|---|
| Yes | ✔ |
| Can't Tell | |
| No | |

HINT: Consider
- what was the goal of the research
- why it was thought important
- its relevance

Comments:

2. Is a qualitative methodology appropriate?

| | |
|---|---|
| Yes | ✔ |
| Can't Tell | |
| No | |

HINT: Consider
- If the research seeks to interpret or illuminate the actions and/or subjective experiences of research participants
- Is qualitative research the right methodology for addressing the research goal

Comments:

#### Is it worth continuing?

3. Was the research design appropriate to address the aims of the research?

| | |
|---|---|
| Yes | ✔ |
| Can't Tell | |
| No | |

HINT: Consider
- if the researcher has justified the research design (e.g. have they discussed how they decided which method to use)

Comments:

CASP
Critical Appraisal Skills Programme

4. Was the recruitment strategy appropriate to the aims of the research?

| | |
|---|---|
| Yes | ✔ |
| Can't Tell | |
| No | |

HINT: Consider
- If the researcher has explained how the participants were selected
- If they explained why the participants they selected were the most appropriate to provide access to the type of knowledge sought by the study
- If there are any discussions around recruitment (e.g. why some people chose not to take part)

Comments:

5. Was the data collected in a way that addressed the research issue?

| | |
|---|---|
| Yes | ✔ |
| Can't Tell | |
| No | |

HINT: Consider
- If the setting for the data collection was justified
- If it is clear how data were collected (e.g. focus group, semi-structured interview etc.)
- If the researcher has justified the methods chosen
- If the researcher has made the methods explicit (e.g. for interview method, is there an indication of how interviews are conducted, or did they use a topic guide)
- If methods were modified during the study. If so, has the researcher explained how and why
- If the form of data is clear (e.g. tape recordings, video material, notes etc.)
- If the researcher has discussed saturation of data

Comments:

CASP
Critical Appraisal
Skills Programme

6. Has the relationship between researcher and participants been adequately considered?

| | |
|---|---|
| Yes | ✔ |
| Can't Tell | |
| No | |

HINT: Consider
- If the researcher critically examined their own role, potential bias and influence during (a) formulation of the research questions (b) data collection, including sample recruitment and choice of location
- How the researcher responded to events during the study and whether they considered the implications of any changes in the research design

Comments: Response Bias: only those who agreed to participate were included, there might be a bias if those who chose not to participate have different experiences or opinions, particularly if they feel less confident or more burdened by the barriers.

#### Section B: What are the results?

7. Have ethical issues been taken into consideration?

| | |
|---|---|
| Yes | ✔ |
| Can't Tell | |
| No | |

HINT: Consider
- If there are sufficient details of how the research was explained to participants for the reader to assess whether ethical standards were maintained
- If the researcher has discussed issues raised by the study (e.g. issues around informed consent or confidentiality or how they have handled the effects of the study on the participants during and after the study)
- If approval has been sought from the ethics committee

Comments: Approval sort from Research Ethics Committee (EC) at the University of Namibia

8. Was the data analysis sufficiently rigorous?

Yes ☐
Can't Tell ☐
No ☐

HINT: Consider
- If there is an in-depth description of the analysis process
- If thematic analysis is used. If so, is it clear how the categories/themes were derived from the data
- Whether the researcher explains how the data presented were selected from the original sample to demonstrate the analysis process
- If sufficient data are presented to support the findings
- To what extent contradictory data are taken into account
- Whether the researcher critically examined their own role, potential bias and influence during analysis and selection of data for presentation

Comments: Could have detailed more clearly the thematic analysis procedures, including: steps taken in the thematic framework analysis, such as coding procedures, how themes emerged, and how the categories were refined.

9. Is there a clear statement of findings?

Yes ☑
Can't Tell ☐
No ☐

HINT: Consider whether
- If the findings are explicit
- If there is adequate discussion of the evidence both for and against the researcher's arguments
- If the researcher has discussed the credibility of their findings (e.g. triangulation, respondent validation, more than one analyst)
- If the findings are discussed in relation to the original research question

Comments:

CASP
Critical Appraisal
Skills Programme

Section C: Will the results help locally?

10. How valuable is the research?

HINT: Consider
- If the researcher discusses the contribution the study makes to existing knowledge or understanding (e.g. do they consider the findings in relation to current practice or policy, or relevant research-based literature
- If they identify new areas where research is necessary
- If the researchers have discussed whether or how the findings can be transferred to other populations or considered other ways the research may be used

Comments: The study addresses a gap in the literature regarding the barriers healthcare workers (HCWs) in Namibia ’ s Zambezi region face in delivering smoking cessation (SC) interventions. findings align with existing literature from other regions, such as Europe, which report similar barriers like insufficient training, lack of patient motivation, and cultural sensitivity regarding SC interventions.
