## Supplementary material for "Efficacy and Best Practices of Health-care worker Smoking Cessation Treatment in Sub-Saharan Africa": Odukoya et al., 2017 CASP checklist

Paper for appraisal and reference: Tobacco Cessation Interventions in Tertiary Hospitals in N

**Section A: Are the results of the study valid?**

1. Did the study address a clearly focused issue?

| | |
|---|---|
| Yes | ☐ |
| Can't Tell | ☐ |
| No | ☑ |

HINT: A question can be 'focused' in terms of
- the population studied
- the risk factors studied
- is it clear whether the study tried to detect a beneficial or harmful effect
- the outcomes considered

Comments: The study focused on assessing the documentation of tobacco cessation interventions by physicians in Nigerian hospitals. The documentation of tobacco use may not truly be representative of the interventions in place.

2. Was the cohort recruited in an acceptable way?

| | |
|---|---|
| Yes | ☐ |
| Can't Tell | ☑ |
| No | ☐ |

HINT: Look for selection bias which might compromise the generalisability of the findings:
- was the cohort representative of a defined population
- was there something special about the cohort
- was everybody included who should have been

Comments: Involved a systematic selection of 1,588 patients from six teaching hospitals across three geopolitical zones in Nigeria, ensuring diversity in the population tested. However, the sample may not be representative of the whole country.

**Is it worth continuing?**

CASP
Critical Appraisal
Skills Programme

3. Was the exposure accurately measured to minimise bias?

Yes ☐
Can't Tell ☑
No ☐

HINT: Look for measurement or classification bias:
- did they use subjective or objective measurements
- do the measurements truly reflect what you want them to (have they been validated)
- were all the subjects classified into exposure groups using the same procedure

Comments: The exposure of physicians' documentation of tobacco use was measured objectively through a uniform checklist applied to patient records. This relied entirely on what was written in patient records, which may not reflect actual practice.

4. Was the outcome accurately measured to minimise bias?

Yes ☐
Can't Tell ☐
No ☑

HINT: Look for measurement or classification bias:
- did they use subjective or objective measurements
- do the measurements truly reflect what you want them to (have they been validated)
- has a reliable system been established for detecting all the cases (for measuring disease occurrence)
- were the measurement methods similar in the different groups
- were the subjects and/or the outcome assessor blinded to exposure (does this matter)

Comments: Outcome (documentation of tobacco cessation activities) was measured using a checklist based on patient records, but the validity of this checklist is not established. Additionally, relying solely on written records for outcome measurement may not be accurate as documenting varies among physicians.

CASP
Critical Appraisal
Skills Programme

5. (a) Have the authors identified all important confounding factors?

Yes ☐
Can't Tell ☑
No ☐

HINT:
- list the ones you think might be important, and ones the author missed

Comments: The author included some confounding factors, such as employment status and tobacco-related diseases. However, other potential confounders, like physician workload, the length of the patient-physician consultation, and the availability of tobacco cessation resources, were not considered.

5. (b) Have they taken account of the confounding factors in the design and/or analysis?

Yes ☑
Can't Tell ☐
No ☐

HINT:
- look for restriction in design, and techniques e.g. modelling, stratified-, regression-, or sensitivity analysis to correct, control or adjust for confounding factors

Comments: Logistic regression was used to adjust for some confounders, such as gender, admission status, and tobacco-related diseases.

6. (a) Was the follow up of subjects complete enough?

Yes ☐
Can't Tell ☑
No ☐

HINT: Consider
- the good or bad effects should have had long enough to reveal themselves
- the persons that are lost to follow-up may have different outcomes than those available for assessment
- in an open or dynamic cohort, was there anything special about the outcome of the people leaving, or the exposure of the people entering the cohort

6. (b) Was the follow up of subjects long enough?

Yes ☐
Can't Tell ☑
No ☐

CASP
Critical Appraisal
Skills Programme

Comments: Not appilcable to this study as this is a one time record audit.

Section B: What are the results?

7. What are the results of this study?

HINT: Consider
- what are the bottom line results
- have they reported the rate or the proportion between the exposed/unexposed, the ratio/rate difference
- how strong is the association between exposure and outcome (RR)
- what is the absolute risk reduction (ARR)

Comments: Results show that only 33.1% of patients had their tobacco use documented, and only 12.9% of identified users received cessation advice.

8. How precise are the results?

HINT:
- look for the range of the confidence intervals, if given

Comments: Can’t tell, as the study provides odds ratios for certain variables but does not include confidence intervals or p-values for all the main outcomes. Therefore, the assessment of the statistical significance of each variable is limited.

9. Do you believe the results?

Yes ☐
Can't Tell ☐
No ☑

HINT: Consider
- big effect is hard to ignore
- can it be due to bias, chance or confounding
- are the design and methods of this study sufficiently flawed to make the results unreliable
- Bradford Hills criteria (e.g. time sequence, dose-response gradient, biological plausibility, consistency)

Comments: The study does not account for the fact that physicians may have provided tobacco cessation advice but failed to document it, leading to an underestimation of the actual rate of intervention.

### Section C: Will the results help locally?

10. Can the results be applied to the local population?

Yes ☑
Can't Tell ☐
No ☐

HINT: Consider whether
- a cohort study was the appropriate method to answer this question
- the subjects covered in this study could be sufficiently different from your population to cause concern
- your local setting is likely to differ much from that of the study
- you can quantify the local benefits and harms

Comments: Similar results may be found in the local regions where the study was carried out, but it may not be generalisable to the rest of the country. The results can influence local smoking cessation policies and the role of physicians in their implementation.

11. Do the results of this study fit with other available evidence?

Yes ☑
Can't Tell ☐
No ☐

Comments: The current findings are consistent with previous research in low-resource settings, where the documentation of tobacco cessation interventions is often poor.

12. What are the implications of this study for practice?

Yes ☑
Can't Tell ☐
No ☐

HINT: Consider
- one observational study rarely provides sufficiently robust evidence to recommend changes to clinical practice or within health policy decision making
- for certain questions, observational studies provide the only evidence
- recommendations from observational studies are always stronger when supported by other evidence

Comments: Yes, the study does provide some important findings that could influence clinical practice in the respective local regions where the data was collected. However, the implications must be interpreted with caution due to the observational nature of the study and the limitations in generalisability.
