## Supplementary material for "Efficacy and Best Practices of Health-care worker Smoking Cessation Treatment in Sub-Saharan Africa": Odukoya, O.O. et al., 2014 CASP checklist

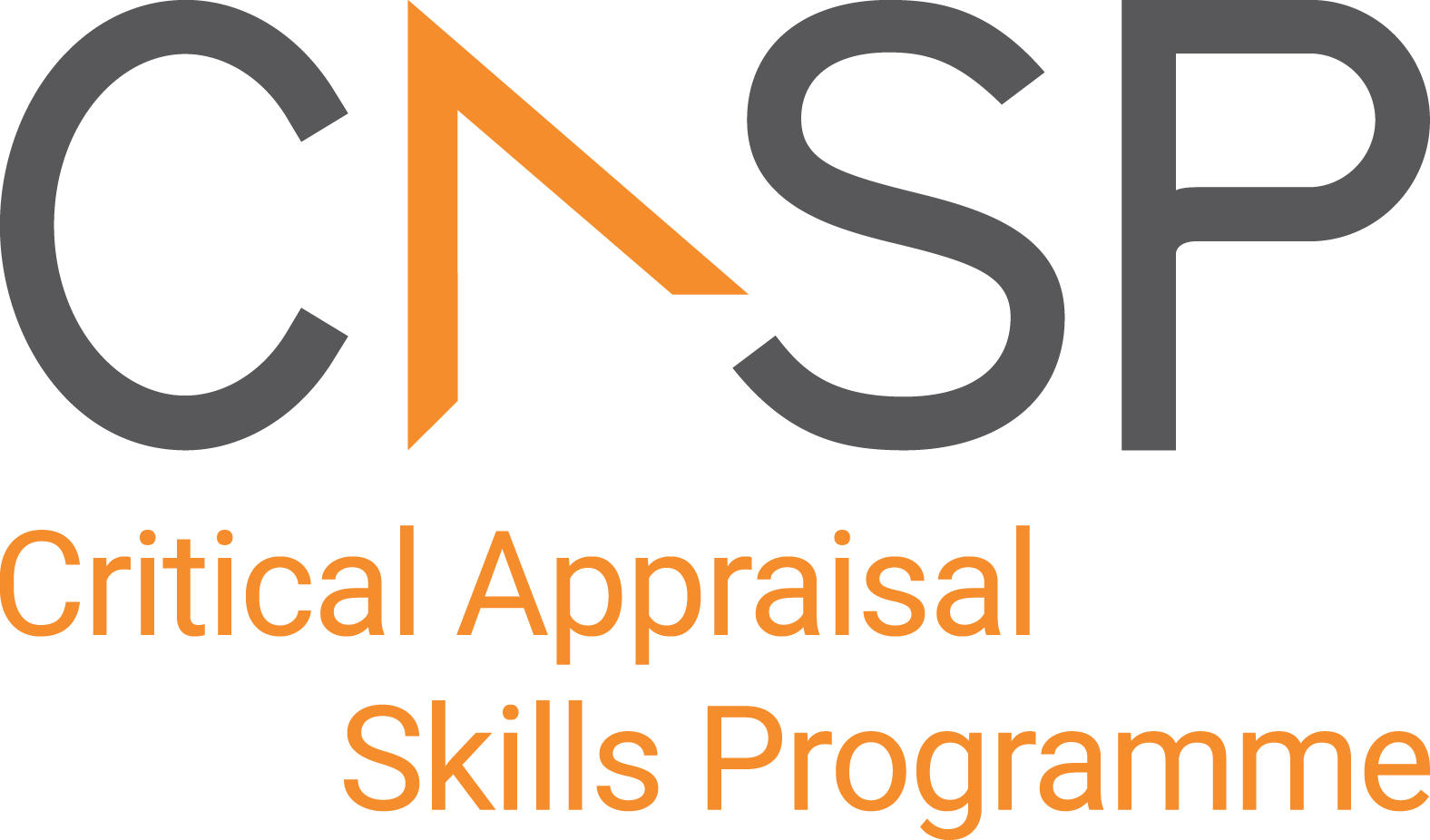
CASP Checklist:

For Cohort Studies

| **Paper Title:** | The effect of a short anti-smoking awareness programme on the knowledge, attitude and practice of cigarette smoking among secondary school students in Lagos state, Nigeria |
| --- | --- |
| **Author:** | Odukoya, O.O. et al., 2014 |
| **Web Link:** | https://www.researchgate.net/publication/264795586_The_effect_of_a_short_anti-smoking_awareness_programme_on_the_knowledge_attitude_and_practice_of_cigarette_smoking_among_secondary_school_students_in_Lagos_State_Nigeria |

| **APPRAISAL SUMMARY**: *List key points from your critical appraisal that need to be considered when assessing the validity of the results and their usefulness in decision-making.* | | |
| --- | --- | --- |
| **Positive/Methodologically sound** | **Negative/Relatively poor methodology** | **Unknowns** |
| • Clear focus on a relevant issue (adolescent smoking).  • Systematic exposure and measurement of outcomes.  • High follow-up rate and consistency in measurement. | • Limited adjustment for potential confounding factors.  • Short follow-up period limits assessment of long-term behaviour change. | Long-term impact of the program on actual smoking behaviour.  Potential for unmeasured confounders influencing results. |

**Referencing recommendation:**

CASP recommends using the Harvard style referencing, which is an author/date method. Sources are cited within the body of your assignment by giving the name of the author(s) followed by the date of publication. All other details about the publication are given in the list of references or bibliography at the end.

Example:

*Critical Appraisal Skills Programme (2024). CASP (insert name of checklist i.e. qualitative studies Checklist.) [online] Available at: insert URL. Accessed: insert date accessed.*

**Creative Commons**

©CASP this work is licensed under the Creative Commons Attribution – Non-Commercial- Share A like. To view a copy of this licence, visit <https://creativecommons.org/licenses/by-nc-sa/4.0/>

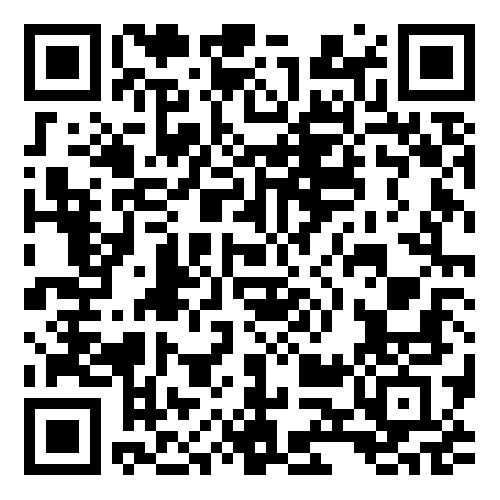
**Need further training on evidence-based decision making?** Our online training courses are helpful for healthcare educational researchers and any other learners who:

- Need to critically appraise and stay abreast of the healthcare research literature as part of their clinical duties.
- Are considering carrying out research & developing their own research projects.
- Make decisions in their role, whether that be policy making or patient facing.

**Benefits of CASP Training:**

- Affordable – courses start from as little as £6
- Professional training – leading experts in critical appraisal training
- Self-directed study – complete each course in your own time
- 12 months access – revisit areas you aren’t sure of and revise
- CPD certification - after each completed module

Scan the QR code below or visit <https://casp-uk.net/critical-appraisal-online-training-courses/> for more information and to start learning more.
