## Supplementary material for "Efficacy and Best Practices of Health-care worker Smoking Cessation Treatment in Sub-Saharan Africa": Shangase et al., 2017 CASP checklist

Paper for appraisal and reference: Barriers to Smoking Cessation Among Drug-Resistant Tub

### Section A: Are the results valid?

1. Was there a clear statement of the aims of the research?

| | |
|---|---|
| Yes | ✔ |
| Can't Tell | |
| No | |

HINT: Consider
- what was the goal of the research
- why it was thought important
- its relevance

Comments: The study had a clear and focused aim - to identify barriers to smoking cessation among drug-resistant tuberculosis patients in South Africa

2. Is a qualitative methodology appropriate?

| | |
|---|---|
| Yes | ✔ |
| Can't Tell | |
| No | |

HINT: Consider
- If the research seeks to interpret or illuminate the actions and/or subjective experiences of research participants
- Is qualitative research the right methodology for addressing the research goal

Comments: A qualitative approach was suitable for exploring perceptions of TB patients regarding smoking cessation.

### Is it worth continuing?

3. Was the research design appropriate to address the aims of the research?

| | |
|---|---|
| Yes | ✔ |
| Can't Tell | |
| No | |

HINT: Consider
- if the researcher has justified the research design (e.g. have they discussed how they decided which method to use)

Comments: The study made use of in-depth interviews, which is appropiate for the aims of this research.

4. Was the recruitment strategy appropriate to the aims of the research?

| | |
|---|---|
| Yes | ✔ |
| Can't Tell | |
| No | |

HINT: Consider
- If the researcher has explained how the participants were selected
- If they explained why the participants they selected were the most appropriate to provide access to the type of knowledge sought by the study
- If there are any discussions around recruitment (e.g. why some people chose not to take part)

Comments: The study selected 20 inpatients with drug-resistant TB who were self-identified smokers, and such participants can provide relevant insights.

5. Was the data collected in a way that addressed the research issue?

| | |
|---|---|
| Yes | ✔ |
| Can't Tell | |
| No | |

HINT: Consider
- If the setting for the data collection was justified
- If it is clear how data were collected (e.g. focus group, semi-structured interview etc.)
- If the researcher has justified the methods chosen
- If the researcher has made the methods explicit (e.g. for interview method, is there an indication of how interviews are conducted, or did they use a topic guide)
- If methods were modified during the study. If so, has the researcher explained how and why
- If the form of data is clear (e.g. tape recordings, video material, notes etc.)
- If the researcher has discussed saturation of data

Comments: In-depth interviews were used to gather information about smoking behaviour, health risk awareness, desire to quit, and barriers to quitting, all of which were relevant to the investigation.

CASP
Critical Appraisal Skills Programme

6. Has the relationship between researcher and participants been adequately considered?

| | |
|---|---|
| Yes | |
| Can't Tell | |
| No | ✔ |

HINT: Consider

- If the researcher critically examined their own role, potential bias and influence during (a) formulation of the research questions (b) data collection, including sample recruitment and choice of location
- How the researcher responded to events during the study and whether they considered the implications of any changes in the research design

Comments: The study does not explicitly discuss the relationship between the researcher and participants.

Section B: What are the results?

7. Have ethical issues been taken into consideration?

| | |
|---|---|
| Yes | ✔ |
| Can't Tell | |
| No | |

HINT: Consider

- If there are sufficient details of how the research was explained to participants for the reader to assess whether ethical standards were maintained
- If the researcher has discussed issues raised by the study (e.g. issues around informed consent or confidentiality or how they have handled the effects of the study on the participants during and after the study)
- If approval has been sought from the ethics committee

Comments: The study has ethical approval by the University of KwaZulu-Natal's Biomedical Research Committee and participants provided individual consent, along with the right to withdraw at any time.

Critical Appraisal Skills Programme

8. Was the data analysis sufficiently rigorous?

| Answer | |
|---|---|
| Yes | ✔ |
| Can't Tell | |
| No | |

HINT: Consider
- If there is an in-depth description of the analysis process
- If thematic analysis is used. If so, is it clear how the categories/themes were derived from the data
- Whether the researcher explains how the data presented were selected from the original sample to demonstrate the analysis process
- If sufficient data are presented to support the findings
- To what extent contradictory data are taken into account
- Whether the researcher critically examined their own role, potential bias and influence during analysis and selection of data for presentation

Comments: Thematic analysis was used to organise points and codes into recurring themes relevant to aim of the study.

9. Is there a clear statement of findings?

| Answer | |
|---|---|
| Yes | ✔ |
| Can't Tell | |
| No | |

HINT: Consider whether
- If the findings are explicit
- If there is adequate discussion of the evidence both for and against the researcher's arguments
- If the researcher has discussed the credibility of their findings (e.g. triangulation, respondent validation, more than one analyst)
- If the findings are discussed in relation to the original research question

Comments: The study clearly presents its findings, identifying key barriers such as nicotine addiction, stress, peer influence, and structural barriers to smoking cessation.

Section C: Will the results help locally?

10. How valuable is the research?

HINT: Consider
- If the researcher discusses the contribution the study makes to existing knowledge or understanding (e.g. do they consider the findings in relation to current practice or policy, or relevant research-based literature
- If they identify new areas where research is necessary
- If the researchers have discussed whether or how the findings can be transferred to other populations or considered other ways the research may be used

Comments: The study provides valuable insights into the specific barriers faced by DR-TB patients in South Africa, which can inform the development of targeted smoking cessation interventions in this context.
