## Supplementary material for "Efficacy and Best Practices of Health-care worker Smoking Cessation Treatment in Sub-Saharan Africa": Bokoro et al., 2020 CASP checklist

Paper for appraisal and reference: Quit attempts among current tobacco users attending the [illegible]

### Section A: Are the results of the study valid?

1. Did the study address a clearly focused issue?

Yes ✔
Can't Tell ☐
No ☐

HINT: A question can be 'focused' in terms of
- the population studied
- the risk factors studied
- is it clear whether the study tried to detect a beneficial or harmful effect
- the outcomes considered

Comments: The study focused on quit attempts among current tobacco users at an outpatient department in South Africa, a clearly defined public health issue.

2. Was the cohort recruited in an acceptable way?

Yes ☐
Can't Tell ✔
No ☐

HINT: Look for selection bias which might compromise the generalisability of the findings:
- was the cohort representative of a defined population
- was there something special about the cohort
- was everybody included who should have been

Comments: The study used consecutive sampling of participants at an outpatient department, which is appropriate for this type of observational research.

Is it worth continuing?

CASP
Critical Appraisal
Skills Programme

3. Was the exposure accurately measured to minimise bias?

| | |
|---|---|
| Yes | ✔ |
| Can't Tell | |
| No | |

HINT: Look for measurement or classification bias:
- did they use subjective or objective measurements
- do the measurements truly reflect what you want them to (have they been validated)
- were all the subjects classified into exposure groups using the same procedure

Comments: Information on tobacco use and quit attempts were assessed using a researcher-administered questionnaire on all subjects. Translators were also provided to those who did not speak English.

4. Was the outcome accurately measured to minimise bias?

| | |
|---|---|
| Yes | |
| Can't Tell | ✔ |
| No | |

HINT: Look for measurement or classification bias:
- did they use subjective or objective measurements
- do the measurements truly reflect what you want them to (have they been validated)
- has a reliable system been established for detecting all the cases (for measuring disease occurrence)
- were the measurement methods similar in the different groups
- were the subjects and/or the outcome assessor blinded to exposure (does this matter)

Comments: Quit attempts as an outcome was self-reported by participants. However bias can be introduced in self-reporting, of which is unavoidable in a study of this nature.

CASP
Critical Appraisal
Skills Programme

5. (a) Have the authors identified all important confounding factors?

Yes ☐
Can't Tell ✔
No ☐

HINT:
- list the ones you think might be important, and ones the author missed

Comments: The study identified some factors associated with quitting, e.g. marital status, but did not comprehensively discuss other potential confounding variables that could affect quit attempts, such as socioeconomic status.

5. (b) Have they taken account of the confounding factors in the design and/or analysis?

Yes ✔
Can't Tell ☐
No ☐

HINT:
- look for restriction in design, and techniques e.g. modelling, stratified-, regression-, or sensitivity analysis to correct, control or adjust for confounding factors

Comments: Patients who were involved in a pilot study were excluded from the final research sample and analysis, to reduce bias.

6. (a) Was the follow up of subjects complete enough?

Yes ☐
Can't Tell ☐
No ✔

HINT: Consider
- the good or bad effects should have had long enough to reveal themselves
- the persons that are lost to follow-up may have different outcomes than those available for assessment
- in an open or dynamic cohort, was there anything special about the outcome of the people leaving, or the exposure of the people entering the cohort

6. (b) Was the follow up of subjects long enough?

Yes ☐
Can't Tell ☐
No ✔

Comments: No follow up as this was a cross-sectional study.

Section B: What are the results?

7. What are the results of this study?

HINT: Consider
- what are the bottom line results
- have they reported the rate or the proportion between the exposed/unexposed, the ratio/rate difference
- how strong is the association between exposure and outcome (RR)
- what is the absolute risk reduction (ARR)

Comments: According to the research, 74% of individuals had tried to quit, with health issues serving as their main driving force. A higher chance of quitting attempts was associated to marital status.

8. How precise are the results?

HINT:
- look for the range of the confidence intervals, if given

Comments: In regression analysis, being married was the only factor independently associated with making a quit attempt ([OR]: 2.13; [CI]: 1.17 – 3.86, p = 0.01).

Critical Appraisal Skills Programme

9. Do you believe the results?

Yes ✔
Can't Tell ☐
No ☐

HINT: Consider
- big effect is hard to ignore
- can it be due to bias, chance or confounding
- are the design and methods of this study sufficiently flawed to make the results unreliable
- Bradford Hills criteria (e.g. time sequence, dose-response gradient, biological plausibility, consistency)

Comments: Despite some limitations, the study provides useful insights into the factors influencing quit attempts among tobacco users in the specific context of South Africa.

### Section C: Will the results help locally?

10. Can the results be applied to the local population?

Yes ✔
Can't Tell ☐
No ☐

HINT: Consider whether
- a cohort study was the appropriate method to answer this question
- the subjects covered in this study could be sufficiently different from your population to cause concern
- your local setting is likely to differ much from that of the study
- you can quantify the local benefits and harms

Comments: The findings are relevant to similar outpatient settings in South Africa and can inform the design of smoking cessation programs in similar contexts.

11. Do the results of this study fit with other available evidence?

Yes ✔
Can't Tell ☐
No ☐

Comments: The findings align with existing evidence on factors influencing smoking cessation, such as the importance of health concerns and social support.

12. What are the implications of this study for practice?

| | |
|---|---|
| Yes | ✔ |
| Can't Tell | |
| No | |

HINT: Consider
- one observational study rarely provides sufficiently robust evidence to recommend changes to clinical practice or within health policy decision making
- for certain questions, observational studies provide the only evidence
- recommendations from observational studies are always stronger when supported by other evidence

Comments: The study suggests that smoking cessation interventions should consider factors such as marital status and health motivations to enhance the effectiveness of quit attempts.
