## Supplementary material for "Efficacy and Best Practices of Health-care worker Smoking Cessation Treatment in Sub-Saharan Africa": Rutebemberwa et al., 2021 CASP checklist

Paper for appraisal and reference: Health workers ’ perceptions on where and how to integra

### Section A: Are the results valid?

1. Was there a clear statement of the aims of the research?

| | |
|---|---|
| Yes | ✔ |
| Can’t Tell | |
| No | |

HINT: Consider
- what was the goal of the research
- why it was thought important
- its relevance

Comments: The aim was clearly stated, to explore the perception of health workers on where and how to integrate tobacco cessation services into TB treatment programs in Uganda.

2. Is a qualitative methodology appropriate?

| | |
|---|---|
| Yes | ✔ |
| Can’t Tell | |
| No | |

HINT: Consider
- If the research seeks to interpret or illuminate the actions and/or subjective experiences of research participants
- Is qualitative research the right methodology for addressing the research goal

Comments: This research seeks to explore patients' perceptions about how and where to introduce smoking cessation services, and so the qualitative methodology is appropriate to explore perceptions and feelings.

### Is it worth continuing?

3. Was the research design appropriate to address the aims of the research?

| | |
|---|---|
| Yes | ✔ |
| Can’t Tell | |
| No | |

HINT: Consider
- if the researcher has justified the research design (e.g. have they discussed how they decided which method to use)

Comments: The study explores why focus group discussions is an appropriat method for this study - althought here aren't explicit mentions about why this method was chosen over others.

CASP
Critical Appraisal
Skills Programme

4. Was the recruitment strategy appropriate to the aims of the research?

| | |
|---|---|
| Yes | |
| Can't Tell | ✔ |
| No | |

HINT: Consider
- If the researcher has explained how the participants were selected
- If they explained why the participants they selected were the most appropriate to provide access to the type of knowledge sought by the study
- If there are any discussions around recruitment (e.g. why some people chose not to take part)

Comments: Participants were purposively selected - the recuritment strategy is well described but not explicitly justified

5. Was the data collected in a way that addressed the research issue?

| | |
|---|---|
| Yes | ✔ |
| Can't Tell | |
| No | |

HINT: Consider
- If the setting for the data collection was justified
- If it is clear how data were collected (e.g. focus group, semi-structured interview etc.)
- If the researcher has justified the methods chosen
- If the researcher has made the methods explicit (e.g. for interview method, is there an indication of how interviews are conducted, or did they use a topic guide)
- If methods were modified during the study. If so, has the researcher explained how and why
- If the form of data is clear (e.g. tape recordings, video material, notes etc.)
- If the researcher has discussed saturation of data

Comments: The data collection strategy was well explained detailing the number of researching experionce, how the focus groups were run, how they were recorded and summarised.

6. Has the relationship between researcher and participants been adequately considered?

| | |
|---|---|
| Yes | |
| Can't Tell | |
| No | ✔ |

HINT: Consider

- If the researcher critically examined their own role, potential bias and influence during (a) formulation of the research questions (b) data collection, including sample recruitment and choice of location
- How the researcher responded to events during the study and whether they considered the implications of any changes in the research design

Comments: The researchers' own positionality, and how it may have affected the data collection strategy and analysis, is not considered.

### Section B: What are the results?

7. Have ethical issues been taken into consideration?

| | |
|---|---|
| Yes | |
| Can't Tell | |
| No | ✔ |

HINT: Consider

- If there are sufficient details of how the research was explained to participants for the reader to assess whether ethical standards were maintained
- If the researcher has discussed issues raised by the study (e.g. issues around informed consent or confidentiality or how they have handled the effects of the study on the participants during and after the study)
- If approval has been sought from the ethics committee

Comments: Details of how ethical standards were maintained, if ethical approval was sought, and details of how the research was explained to participants are not present in the paper.

8. Was the data analysis sufficiently rigorous?

| | |
|---|---|
| Yes | ✔ |
| Can't Tell | |
| No | |

HINT: Consider
- If there is an in-depth description of the analysis process
- If thematic analysis is used. If so, is it clear how the categories/themes were derived from the data
- Whether the researcher explains how the data presented were selected from the original sample to demonstrate the analysis process
- If sufficient data are presented to support the findings
- To what extent contradictory data are taken into account
- Whether the researcher critically examined their own role, potential bias and influence during analysis and selection of data for presentation

Comments: How the analysis was conducted is detailed as well as how disagreements were resolved.

9. Is there a clear statement of findings?

| | |
|---|---|
| Yes | ✔ |
| Can't Tell | |
| No | |

HINT: Consider whether
- If the findings are explicit
- If there is adequate discussion of the evidence both for and against the researcher's arguments
- If the researcher has discussed the credibility of their findings (e.g. triangulation, respondent validation, more than one analyst)
- If the findings are discussed in relation to the original research question

Comments: The results are explicitly stated and each idea explored individually with quotations to evidence the results.

CASP
Critical Appraisal
Skills Programme

Section C: Will the results help locally?

10. How valuable is the research?

HINT: Consider
- If the researcher discusses the contribution the study makes to existing knowledge or understanding (e.g. do they consider the findings in relation to current practice or policy, or relevant research-based literature
- If they identify new areas where research is necessary
- If the researchers have discussed whether or how the findings can be transferred to other populations or considered other ways the research may be used

Comments: The research frames the findings in the context of the existing literature and it makes recommendations for policy, practice and health education that would allow for tobacco cessation to be integrated into TB management.
