## Supplementary material for "Efficacy and Best Practices of Health-care worker Smoking Cessation Treatment in Sub-Saharan Africa": Thirlway et al., 2021 CASP checklist

Paper for appraisal and reference: obacco use and cessation in the context of ART adherence

### Section A: Are the results valid?

1. Was there a clear statement of the aims of the research?

Yes ✔
Can't Tell
No

HINT: Consider
- what was the goal of the research
- why it was thought important
- its relevance

Comments: The aim of this study is clearly stated at the end of the introduction: "to explore patterns of tobacco use and cessation and facilitators and barriers to cessation among adults receiving treatment for HIV in Uganda, with a view to developing effective smoking cessation interventions"

2. Is a qualitative methodology appropriate?

Yes ✔
Can't Tell
No

HINT: Consider
- If the research seeks to interpret or illuminate the actions and/or subjective experiences of research participants
- Is qualitative research the right methodology for addressing the research goal

Comments: The research seeks to explore barriers and facilitators to cessation among adults receiving HIV treatment which the qualitative method of semi-structured interview facilitates.

### Is it worth continuing?

3. Was the research design appropriate to address the aims of the research?

Yes
Can't Tell ✔
No

HINT: Consider
- if the researcher has justified the research design (e.g. have they discussed how they decided which method to use)

Comments: Semi structured interviews with participants was an appropriate design to address the research question and aims, however, it was not explicitly justified.

4. Was the recruitment strategy appropriate to the aims of the research?

| | |
|---|---|
| Yes | |
| Can't Tell | ✔ |
| No | |

HINT: Consider
- If the researcher has explained how the participants were selected
- If they explained why the participants they selected were the most appropriate to provide access to the type of knowledge sought by the study
- If there are any discussions around recruitment (e.g. why some people chose not to take part)

Comments: Convenience sampling was used to recruit participants, with the help of clinical staff, from attending HIV clinics in areas known to have high rates of tobacco use. However the recruitment strategy wasn't explicitly justified.

5. Was the data collected in a way that addressed the research issue?

| | |
|---|---|
| Yes | ✔ |
| Can't Tell | |
| No | |

HINT: Consider
- If the setting for the data collection was justified
- If it is clear how data were collected (e.g. focus group, semi-structured interview etc.)
- If the researcher has justified the methods chosen
- If the researcher has made the methods explicit (e.g. for interview method, is there an indication of how interviews are conducted, or did they use a topic guide)
- If methods were modified during the study. If so, has the researcher explained how and why
- If the form of data is clear (e.g. tape recordings, video material, notes etc.)
- If the researcher has discussed saturation of data

Comments: The data was collected via semi-structured interview and were audio recorded and supplemented with additional data such as interviewers' notes.

CASP
Critical Appraisal
Skills Programme

6. Has the relationship between researcher and participants been adequately considered?

Yes ☐
Can't Tell ☐
No ☑

HINT: Consider
- If the researcher critically examined their own role, potential bias and influence during (a) formulation of the research questions (b) data collection, including sample recruitment and choice of location
- How the researcher responded to events during the study and whether they considered the implications of any changes in the research design

Comments: The researcher don't directly consider their position in relation to the participants beyond using health workers to help recruit participants to bridge social barriers.

### Section B: What are the results?

7. Have ethical issues been taken into consideration?

Yes ☐
Can't Tell ☑
No ☐

HINT: Consider
- If there are sufficient details of how the research was explained to participants for the reader to assess whether ethical standards were maintained
- If the researcher has discussed issues raised by the study (e.g. issues around informed consent or confidentiality or how they have handled the effects of the study on the participants during and after the study)
- If approval has been sought from the ethics committee

Comments: The study discusses the way that confidentiality was maintained (e.g., socioeconomic status and participant locations omitted) and formal ethical approval was thought. However, there's no mention of how informed consent was ensured or consideration for other ethical concerns relating to the potential effects of the study.

CASP
Critical Appraisal Skills Programme

8. Was the data analysis sufficiently rigorous?

Yes [ ]
Can't Tell [✔]
No [ ]

HINT: Consider
- If there is an in-depth description of the analysis process
- If thematic analysis is used. If so, is it clear how the categories/themes were derived from the data
- Whether the researcher explains how the data presented were selected from the original sample to demonstrate the analysis process
- If sufficient data are presented to support the findings
- To what extent contradictory data are taken into account
- Whether the researcher critically examined their own role, potential bias and influence during analysis and selection of data for presentation

Comments: The description of the analysis process is very bare - no explanation of how themes were derived, how contradictory data was taken into account or consideration of positionality

9. Is there a clear statement of findings?

Yes [✔]
Can't Tell [ ]
No [ ]

HINT: Consider whether
- If the findings are explicit
- If there is adequate discussion of the evidence both for and against the researcher's arguments
- If the researcher has discussed the credibility of their findings (e.g. triangulation, respondent validation, more than one analyst)
- If the findings are discussed in relation to the original research question

Comments: Findings are explicitly mentioned and explored.

Critical Appraisal Skills Programme

Section C: Will the results help locally?

10. How valuable is the research?

HINT: Consider
- If the researcher discusses the contribution the study makes to existing knowledge or understanding (e.g. do they consider the findings in relation to current practice or policy, or relevant research-based literature
- If they identify new areas where research is necessary
- If the researchers have discussed whether or how the findings can be transferred to other populations or considered other ways the research may be used

Comments: The researchers consider how this study could influence policy to support people living with HIV giving up tobacco in centering patient care, and appropriate recommendations are made.
