## Supplementary material for "Efficacy and Best Practices of Health-care worker Smoking Cessation Treatment in Sub-Saharan Africa": Louwagie, G.M. et al., 2022 CASP checklist

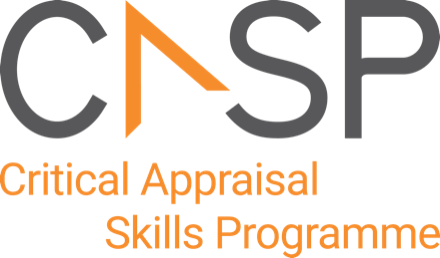

**CASP Randomised Controlled Trial Standard Checklist:**

11 questions to help you make sense of a randomised controlled trial (RCT)

**Main issues for consideration:** Several aspects need to be considered when appraising a randomised controlled trial:

Is the basic study design valid for a randomised controlled trial? (Section A)

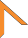

Was the study methodologically sound? (Section B)

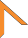

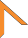
 What are the results? (Section C)

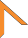
 Will the results help locally? (Section D)

The 11 questions in the checklist are designed to help you think about these aspects systematically.

**How to use this appraisal tool:** The first three questions (Section A) are screening questions about the validity of the basic study design and can be answered quickly. If, in light of your responses to Section A, you think the study design is valid, continue to Section B to assess whether the study was methodologically sound and if it is worth continuing with the appraisal by answering the remaining questions in Sections C and D.

Record ‘Yes’, ‘No’ or ‘Can’t tell’ in response to the questions. Prompts below all but one of the questions highlight the issues it is important to consider. Record the reasons for your answers in the space provided. As CASP checklists were designed to be used as educational/teaching tools in a workshop setting, we do not recommend using a scoring system.

**About CASP Checklists:** The CASP RCT checklist was originally based on JAMA Users’ guides to the medical literature 1994 (adapted from Guyatt GH, Sackett DL and Cook DJ), and piloted with healthcare practitioners. This version has been updated taking into account the CONSORT 2010 guideline (<http://www.consort-statement.org/consort-2010>, accessed 16 September 2020).

**Citation:** CASP recommends using the Harvard style, i.e., *Critical Appraisal Skills Programme (2021). CASP (insert name of checklist i.e. Randomised Controlled Trial) Checklist. [online] Available at: insert URL. Accessed: insert date accessed.*

©CASP this work is licensed under the Creative Commons Attribution – Non-Commercial- Share A like. To view a copy of this licence, visit <https://creativecommons.org/licenses/by-sa/4.0/>

Critical Appraisal Skills Programme (CASP) [www.casp-uk.net](http://www.casp-uk.net/) Part of OAP Ltd

**Study and citation:** Effect of a brief motivational interview and text message intervention targeting tobacco smoking, alcohol use and medication adherence to improve tuberculosis treatment outcomes in adult patients with tuberculosis: a multicentre, randomised controlled trial of the ProLife programme in South Africa (Louwagie et al., 2022)

**Section D: Will the results help locally?**

| **10**. | **Can the results be applied to your local population/in your context?**  *CONSIDER:*   - *Are the study participants similar to the people in your care?* - *Would any differences between your population and the study participants alter the outcomes reported in the study?* - *Are the outcomes important to your population?* - *Are there any outcomes you would have wanted information on that have not been studied or reported?* - *Are there any limitations of the study that would affect your decision?* | Yes No Can’t tell  🞏 🞏 🞏 |
| --- | --- | --- |
| **11.** | **Would the experimental intervention provide greater value to the people in your care than any of the existing interventions?**  *CONSIDER:*   - *What resources are needed to introduce this intervention taking into account time, finances, and skills development or training needs?* - *Are you able to disinvest resources in one or more existing interventions in order to be able to re-invest in the new intervention?* | Yes No Can’t tell  🞏 🞏 🞏 |
| **APPRAISAL SUMMARY:** *Record key points from your critical appraisal in this box. What is your conclusion about the paper? Would you use it to change your practice or to recommend changes to care/interventions used by your organisation? Could you judiciously implement this intervention without delay?*  This study comprised a well-designed randomised controlled trial and comprehensive presentation of primary and secondary outcomes. It demonstrated that MI by health workers was effective in promoting smoking cessation among tuberculosis patients in South Africa, with smoking cessation rates approximately doubling. Although participants and healthcare workers were not blinded, and there was no cost-effectiveness data provided, the intervention could be beneficial in healthcare settings with limited resources, as it frees up nurse time for other critical tasks. | | |
